# Post-vaccination chemosensory outcomes in COVID-19-associated dysfunction

**DOI:** 10.1101/2025.05.28.25328470

**Authors:** Sachiko Koyama, Carla Mucignat-Caretta, Maria A. Klyuchnikova, Tatiana K. Laktionova, Svetlana Kopishinskaia, Ilya G. Kvasha, Junpeng Ye, Elena Cantone, Shima Parsa, Maria Dolors Guàrdia, Valentin Hivert, Solal Chauquet, Jingguo Chen, Giulio Centorame, Valentina Parma, Kathrin Ohla, Vera V. Voznessenskaya, Liang-Dar Hwang

**Affiliations:** Department of Medicine, Indiana University School of Medicine, Indianapolis, Indiana, USA; Department of Molecular Medicine, University of Padova, Padova, Italy; Severtsov Institute of Ecology & Evolution, Russian Academy of Sciences, Moscow, Russia; Centre de Recherche du CHU de Québec, Laval University, Quebec, Canada; Institute for Molecular Bioscience, The University of Queensland, Brisbane, Queensland, Australia; Department of Neuroscience, Reproductive and Dentistry Science, ENT Unit, Federico II University of Naples, Naples, Italy; Shiraz Institute for Stem Cell & Regenerative Medicine, Shiraz University of Medical Sciences, Shiraz, Iran; Food Quality and Technology, IRTA-Food Research Institute, Monells, Spain; The Department of Otolaryngology Head and Neck Surgery, Second Affiliated Hospital of Xi’an Jiaotong University, Xi’an, China; Monell Chemical Senses Center, Philadelphia, Pennsylvania, USA; Science & Research, dsm-firmenich, Satigny, Switzerland

**Keywords:** COVID-19, vaccines, olfactory disorders, parosmia, phantosmia, post-acute COVID-19 syndrome

## Abstract

Olfactory and taste dysfunction are common symptoms of COVID-19 and post-acute COVID-19 syndrome, yet the effects of COVID-19 vaccines on chemosensory perception remain incompletely understood. This global, multilingual, online survey assessed post-vaccination changes in chemosensory function among individuals with and without COVID-19-related chemosensory impairment. Between May 2022 and August 2023, 2,955 responses were collected via convenience sampling, of which 1,352 were included in the analyses. Participants reported vaccination status, side effects, and chemosensory function before and after each vaccine dose. Pfizer-BioNTech accounted for 46.2% of doses, followed by Sputnik V (16.3%), Moderna (15.4%), AstraZeneca (8.9%), and Sinopharm (7.4%). More than 90% of participants reported no change in their general sense of smell or taste following vaccination, regardless of pre-existing chemosensory impairment. Among participants with qualitative chemosensory distortions (one-third of the sample), improvement was reported by 11-18% for parosmia, 20-29% for phantosmia, and 12-21% for taste distortion, depending on the vaccine dose, while worsening was reported by 3% or fewer. Side effects varied by vaccine type and were more frequent among individuals with worsened chemosensory symptoms. These findings suggest that COVID-19 vaccination is unlikely to adversely affect chemosensory function for most individuals. Given the observational design and reliance on self-reported data, the results should be interpreted cautiously. Future longitudinal studies using objective measures are needed to clarify these associations.

## INTRODUCTION

Following the emergence of COVID-19 (Coronavirus disease 2019), caused by SARS-CoV-2, unprecedented global efforts led to the rapid development of effective vaccines. Within one year of the initial outbreak, multiple COVID-19 vaccines were authorized and deployed worldwide [1–3]. SARS-CoV-2 vaccines can be broadly classified into four types based on their antigen delivery mechanisms: mRNA vaccines, adenovirus vector vaccines, inactivated virus vaccines, and recombinant protein vaccines (see reviews [1–3] and the World Health Organization overview [4].

Since the onset of the pandemic, numerous studies have characterized the clinical manifestations of COVID-19 [5,6]. Acute infections can present with a wide range of symptoms, including fever, cough, fatigue, shortness of breath, gastrointestinal and neurological symptoms, as well as chemosensory dysfunction. Asymptomatic infections have also been reported [7,8]. Recovery trajectories vary considerably between individuals [9–12], with symptoms persisting for three months or longer after infection being commonly referred to as post-COVID syndrome, post-acute sequelae of COVID-19, or Long COVID [13,14]. This later became a Medical Subject Headings (MeSH) term, “Post-acute COVID-19 syndrome” (PCS). Although prevalence estimates vary due to the heterogeneous and non-specific nature of symptoms, recent studies suggest that PCS may affect up to 45% of individuals following SARS-CoV-2 infection [13,14].

Chemosensory dysfunction, encompassing impaired or distorted smell, taste, or chemesthetic perception, is a well-established feature of both acute COVID-19 and PCS [15–20]. Although chemosensory dysfunctions were previously observed with other viral infections of the upper respiratory system, their occurrence was relatively rare compared to COVID-19 [21], in which up to 77% of COVID-19 patients reported smell dysfunction and 44% reported taste dysfunction [15,20,22–24].

Soon after COVID-19 vaccines became widely available, changes in chemosensory perception following vaccination were reported, including both improvement and worsening of symptoms, primarily in case reports and cohort studies. For example, a large U.S.-based study using electronic medical records from more than 96 million individuals reported extremely low incidence rates of post-vaccination smell (0.003%) and/or taste (0.009%) disturbance [25]. Another UK-based cohort study of individuals with PCS reported reduced odds of loss of smell (-12.5%) and loss of taste (-9.2%) after receiving adenovirus vector or mRNA vaccines [26]. Temporary olfactory dysfunction following vaccination has also been reported in isolated cases [27–29]. Although vaccine types and brands differ in their side-effect profiles [30], the relationship between COVID-19 vaccination and changes in chemosensory perception has not been systematically examined across diverse populations.

To address this gap, we conducted a large global online survey to investigate the relationship between COVID-19 vaccination and post-vaccination changes in chemosensory perception among individuals with and without COVID-19 related chemosensory impairment. We examined vaccination patterns across countries, changes in the general ability to smell and taste, and qualitative chemosensory distortion, as well as their relationships with vaccine-related side effects.

## MATERIALS AND METHODS

### Survey design

The survey started with collecting the date and brand of each COVID-19 vaccine received, followed by information on post-vaccination side effects. Participants selected side effects from a predefined list (including fever, headache, chills, diarrhea, vomit, stomach ache, joint ache, cough, sore throat, shortness of breath, muscle soreness, lightheadedness, dizziness, nausea, fatigue, nasal congestion, burning nose, heart problems, sleep disturbances, blood clots, skin rash, menstrual changes, brain fog, and others) and reported the duration of the side effects.

Given the focus on chemosensory outcomes, the survey included detailed questions on changes in smell, taste, and chemesthesis. Participants reported whether each modality had been affected by COVID-19 before vaccination, whether chemosensory dysfunction improved, worsened, or remained unchanged after vaccination, as well as the time of onset, and the duration of the changes. Participants also reported experiencing parosmia, phantosmia, and distorted taste. All chemosensory questions were answered separately for each vaccine dose received.

The survey was deployed in English, Chinese (traditional and simplified), French, German, Italian, Japanese, Persian, Russian, and Spanish via the online platform Qualtrics hosted at Indiana University (see **Supplementary Materials** for the full survey in English). The study was approved by the Global Consortium for Chemosensory Research (GCCR) Study Selection Team (ID: NDS006), the Institute Review Board of Indiana University (Protocol #: 15050), the Human Research Ethics Committee of the University of Queensland (Project ID: 2022/HE000839), and the Bioethics Committee at the A.N. Severtsov Institute of Ecology & Evolution of Russian Academy of Sciences (Protocol #.2022-63). The study was pre-registered on the Open Science Framework (OSF) [31].

### Data collection and preprocessing

The online crowdsourced survey was distributed via social media, personal communication, GCCR newsletters (over 700 subscribers), and emails sent to over 6000 participants of prior GCCR studies [15,16] who had consented to be recontacted. A total of 2,955 responses were received between May 2022 and August 2023.

Open-text responses from non-English surveys were translated into English and then merged for analysis. Data preprocessing was conducted in accordance with the pre-registered exclusion criteria. A total of 1,602 respondents were excluded because they did not review all questions before exiting the survey (n = 1,066), reported pre-existing health conditions (n = 228), did not receive a COVID-19 vaccine (n = 206), were pregnant after the outbreak of COVID (a known modulator of chemosensory ability [32,33], n = 69) or younger than 18 years old (n = 33) (**Supplementary Figure 1**). Participants with pre-existing health conditions were those who selected any of the following: “Neurological pathologies”, “Psychiatric pathologies”, “Head and neck trauma”, “Nasal sinus surgery”, “Head and neck radiation”, and “Impaired smell and taste perception” to the question “*Do you have below pre-existing conditions before COVID and/or before vaccination?*” One additional respondent was excluded due to non-compliance with study instructions. The final sample of 1,352 participants was included in the analyses.

Participants were required to answer questions related to vaccination status, while other items were optional, resulting in varying sample sizes across analyses.

### Data analysis

Sample characteristics were summarized by age, sex, country of origin, vaccination status (number of vaccination doses and boosters), vaccine brand (counts and percentages), and pre-vaccination chemosensory status, *i.e.*, whether chemosensory perception was affected by COVID-19 before vaccination. Post-vaccination chemosensory outcomes (improvement, worsening, or no change) were reported for the full sample and stratified by the presence or absence of COVID-19-related chemosensory impairment prior to vaccination. Pre-existing chemosensory dysfunction was defined as unresolved COVID-19-related smell or taste loss, or the presence of parosmia, phantosmia, or distorted taste before vaccination.

Proportions of post-vaccination chemosensory changes were compared across vaccine brands. Given country-level differences in vaccine availability, country- and region-specific summaries (global regions and European regions) were also provided. Differences in chemosensory outcomes across vaccine brands were assessed using Fisher’s test of homogeneity. Binomial tests were used to compare the proportion of participants reporting improvement to the proportion reporting worsening for each vaccine brand. To evaluate potential recall bias, follow-up durations (time between the vaccination date and survey completion) were compared across outcome groups using one-way analysis of variance (ANOVA).

For vaccine side effects, the frequency of each symptom was first summarized by vaccine dose. Vaccine brands were grouped into four types: mRNA vaccines, adenovirus vector vaccines, inactivated virus vaccines, and recombinant protein vaccines (see **Supplementary Note**). Incidence rates of side effects were calculated across the first three doses of vaccines within each type. Within each vaccine type, differences in side-effect incidence across doses were evaluated using means of the k-proportions test with the Marascuilo Procedure. Lastly, the total number of reported side effects was compared across groups defined by post-vaccination changes in smell and taste using one-way ANOVA and pairwise t-tests.

All other analyses were performed in R (version 4.3.0) and RStudio (version 2023.03.0 Build 386), except for the k-proportions tests, which were performed using XLSTAT (version 2021.1.1).

## RESULTS

### Sample characteristics

The final analytic sample comprised 1,352 participants from 52 countries (**Supplementary Table 1)**. Most participants were aged between 25 and 64 years (71.6%), and 46.99% identified as cis women (**Supplementary Figure 2**). Pre-vaccination chemosensory impairments due to COVID-19 were reported by 599 of 1220 participants with complete taste and smell data (49%). Of these, 416 reported impairments in both smell and taste, and most of the remainder reported smell impairment only (**Supplementary Figure 3**). Pre-vaccination parosmia, phantosmia, and distorted taste were reported by 424, 265, and 380 participants, respectively (**Supplementary Tables 2 & 3)**. Country-specific prevalence estimates are provided in **Supplementary Table 4**.

Approximately half of the participants received two vaccine doses plus one booster (46.1%), 28.2% received two doses plus two boosters, 20% received two doses only, and 5.8% received a single dose (see **Figure 1A** for the country-specific vaccination status). Among the 1,200 participants who received at least two doses, two doses of Pfizer-BioNTech was the most common combination (44.2%, n = 531), except in Russia, Kazakhstan, Iran, the UK, and Switzerland, where Sputnik V, Sinopharm, Astra-Zeneca, or Moderna predominated (**Figure 1B**). Among the 824 participants who received at least two doses and one booster, Pfizer-BioNTech double doses were most often combined with a booster from the same brand (n = 276, 33.5%) or with a booster from Moderna (n = 109, 13.2%). The third most common combination was two doses of Sputnik V plus a booster of Sputnik Light (n = 59, 7.2%, **Supplementary Table 5**). Overall, Pfizer-BioNTech accounted for 46.2% of all 3634 vaccine doses administered, followed by Sputnik V (16.3%), Moderna (15.4%), AstraZeneca (8.9%), and Sinopharm (7.4%).

**Figure 1.**
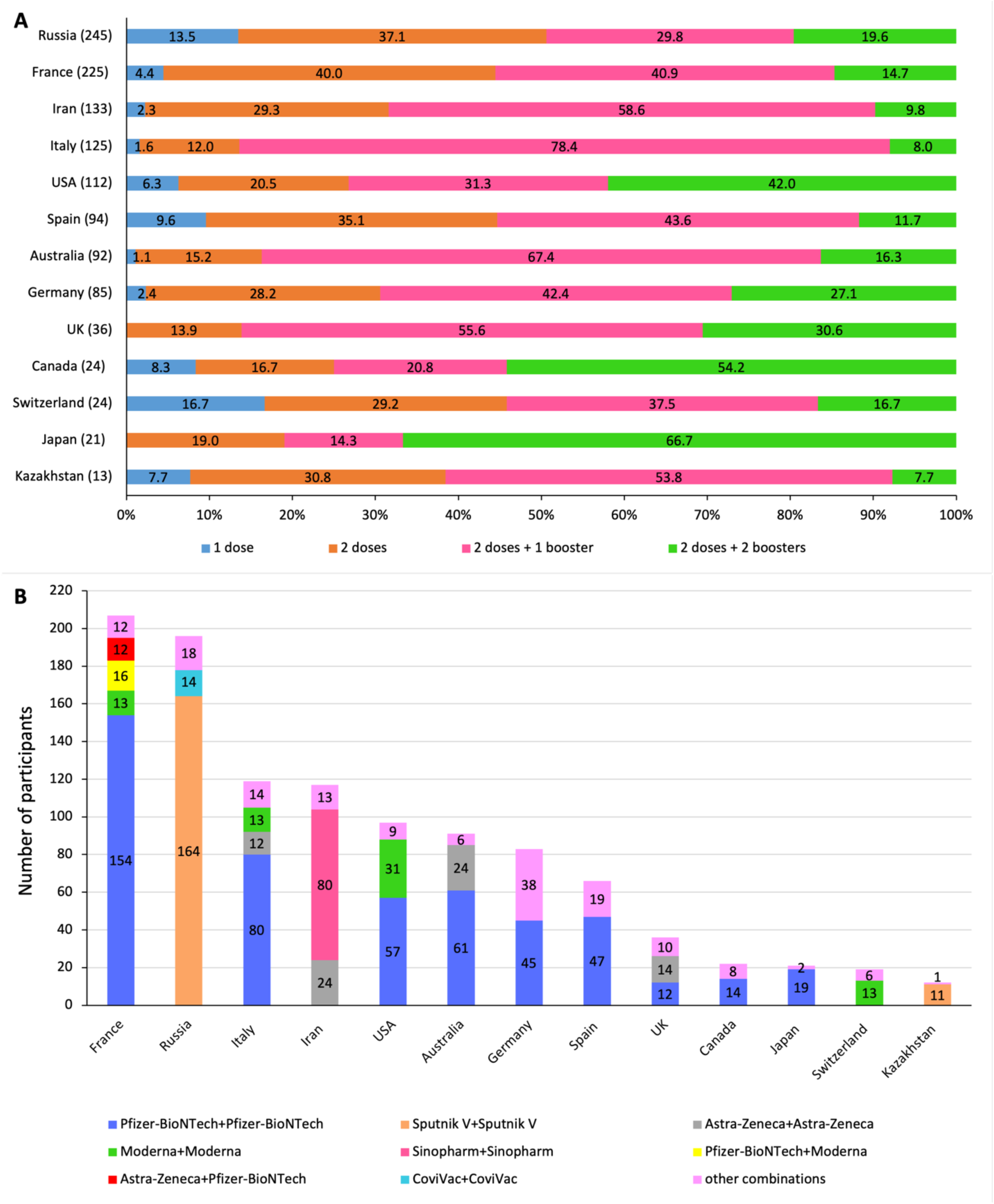
Distribution of vaccination status of participants (A) and combinations of the first two doses received (B) in different countries. Only countries with ≥10 respondents were included. The number in brackets represents the total number of vaccinated participants per country.

### Changes in chemosensory perception

Among all participants, 94% or more reported no change in their general ability to smell or taste perception after the first vaccine dose. When stratified by pre-existing COVID-19-related chemosensory impairment, 90% participants with impairment and 97% without impairment reported no changes (**Supplementary Figure 4**; **Supplementary Table 6**). Among participants with pre-existing impairments, 3-8% reported improvement and 2-7% reported worsening in smell or taste, depending on dose number (**Figure 2**).

**Figure 2.**
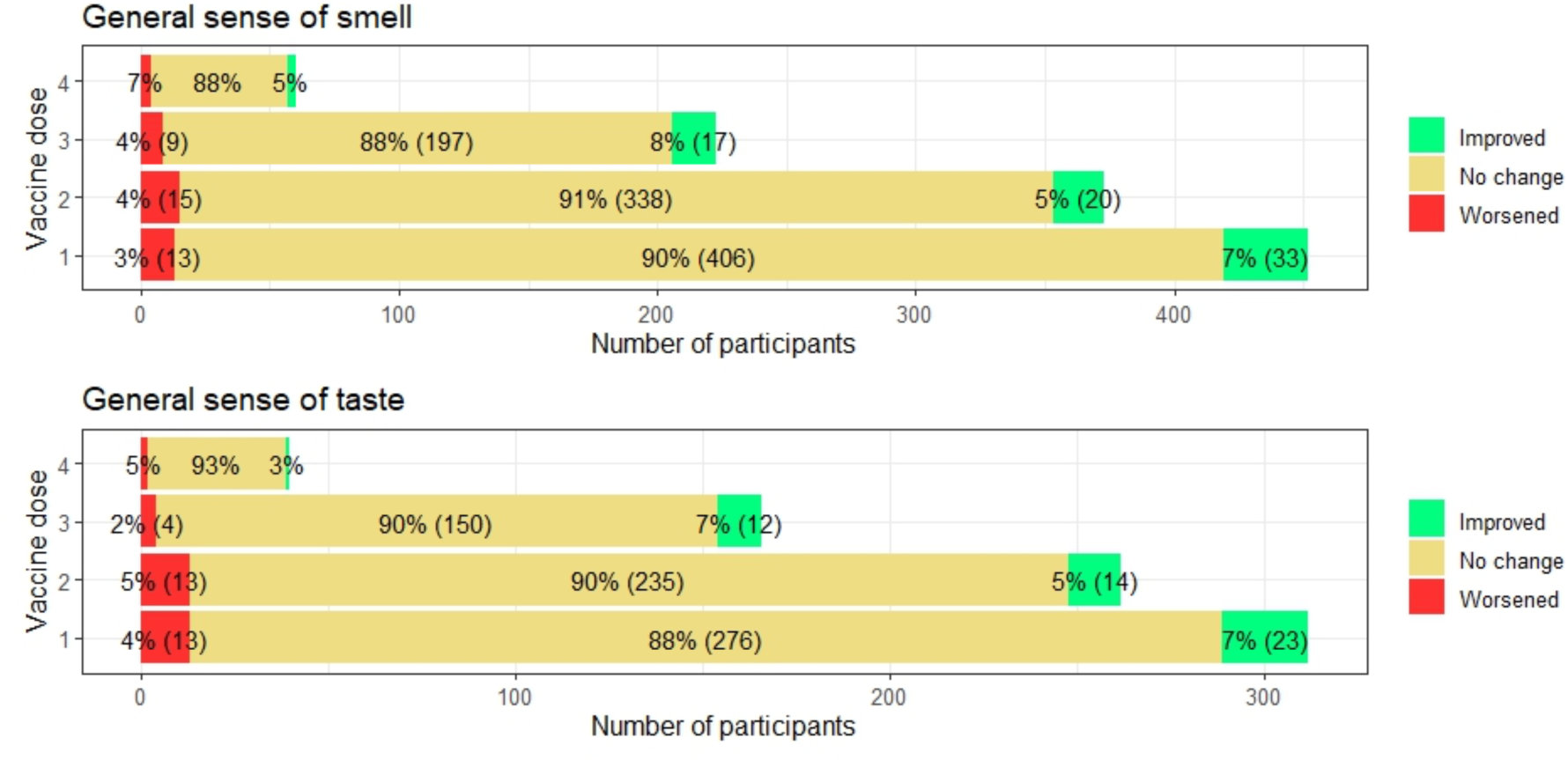
Changes in the senses of smell and taste after each dose of vaccine in participants with pre-existing chemosensory impairment due to COVID-19. The number of participants per condition for the first three doses is shown in parentheses.

For qualitative chemosensory symptoms, 66% or more of participants did not report parosmia, phantosmia, or distorted taste before or after vaccination, and 2% or fewer reported new onset following vaccination (**Supplementary Tables 2 & 3**). Among participants with pre-existing qualitative impairments, improvement was reported by 11-18% for parosmia, 20-29% for phantosmia, and 12-21% for distorted taste, depending on dose, while worsening was reported by 3% or fewer (**Figure 3**).

**Figure 3.**
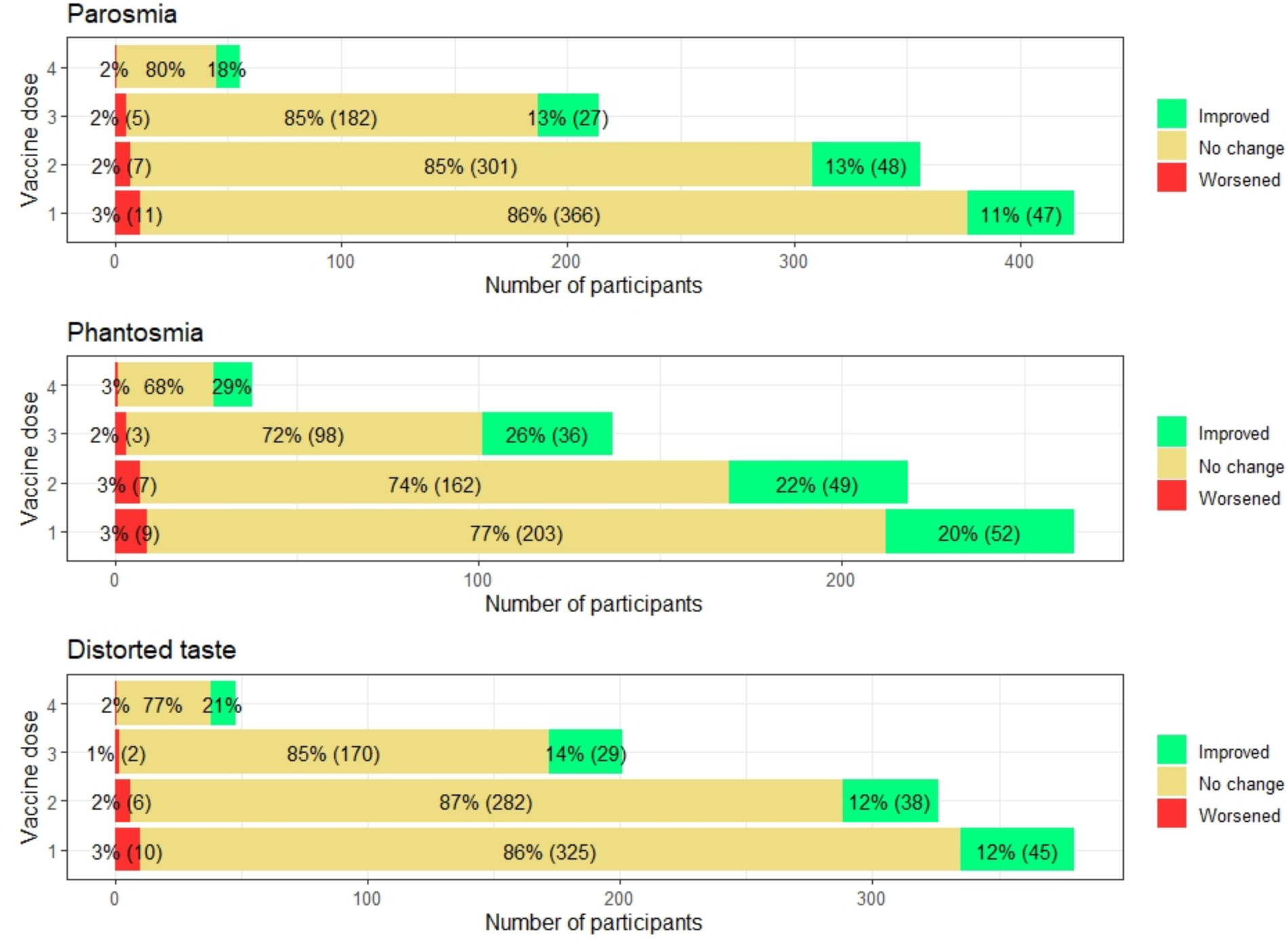
Changes in parosmia/phantosmia/distorted taste after each dose of vaccine in participants with pre-existing sensory impairment due to COVID-19. The number of participants per condition for the first three doses is shown in parentheses.

Among the 75 participants who reported the timing and duration of post-vaccination chemosensory changes, approximately two-thirds indicated that the onset occurred on the day following vaccination, and most reported symptom duration longer than one month (**Supplementary Tables 2 and 3**). The time elapsed between the first vaccine dose and survey completion did not differ across participants reporting improvement, worsening, or no change (∼20 months, p > 0.05; **Supplementary Table 7**).

### Changes in chemosensory perception by vaccine brands

Among all participants, post-vaccination improvement in general smell after the first dose was most frequently reported following Moderna (9.6%, 95% CI [4.4, 14.8]) and least frequently following Sputnik V (0.8%, 95% CI [-0.3, 1.9]). Worsening was most frequent following Sputnik V (4.5%, 95% CI [1.9, 7.1]) and least frequent following Pfizer-BioNTech (1.3%, 95% CI [0.4, 2.3]) (**Supplementary Table 8**). Among participants with pre-existing smell dysfunctions (n = 432), improvement remained most frequent following Moderna (8 of 67, 11.9 %, 95% CI [4.2, 19.7]), while worsening was most frequent following Sputnik V (3 of 52, 5.8%, 95% CI [0.0, 12.1]). Similar patterns were observed after two doses of the same vaccines. Country- and region-specific summaries are provided in **Supplementary Table 9**. Post-vaccination changes in taste followed similar patterns (**Supplementary Tables 10 & 11**).

Although overall improvement rates differed across vaccine brands (p < 0.05), there was no evidence of systematic differences between improvement and worsening rates within individual brands. Given the small and unequal sizes of vaccine groups, these results should be interpreted with caution.

Among individuals with pre-existing parosmia, improvement after the first dose was most frequent following Moderna (24.5%, 95% CI [12.9, 36.1]) and least frequent following Astra-Zeneca (6.7%, 95% CI [-0.6, 14.0]) (**Table 1**). For phantosmia, improvement was the highest following Moderna (34.4%, 95% CI [17.9, 50.8]) and lowest following Pfizer-BioNTech (17.1%, 95% CI [11.1, 23.1]) (**Table 2**). For distorted taste, Sputnik showed the highest proportions of both improvements (20.0%, 95% CI [6.7, 33.3]) and worsening (8.6%, 95% CI [-0.7, 17.8]) (**Table 3**). Overall, improvement was more frequent than worsening across qualitative chemosensory outcomes (p < 0.001), with no significant differences across brands, except for parosmia after the first dose. Country- and region-specific results are summarized in **Supplementary Tables 12-14**.

**Table 1.** Post-vaccination changes in pre-existing parosmia by vaccine brands.

| After the first dose of vaccine |  |  |  |  |  |  |
| --- | --- | --- | --- | --- | --- | --- |
| Brands | Vaccinated participants | Improved parosmia |  | Worsened parosmia |  | Improved vs worsened<sup>d</sup> |
|  | N | N | Proportion | N | Proportion | P |
| **Pfizer-BioNTech<sup>a</sup>** | 224 | 20 | 8.9% | 4 | 1.8% | 0.002 |
| **Moderna<sup>a</sup>** | 53 | 13 | 24.5% | 0 | 0% | <0.001 |
| **Sputnik V<sup>b</sup>** | 53 | 8 | 15.1% | 4 | 7.5% | 0.39 |
| **Astra-Zeneca<sup>b</sup>** | 45 | 3 | 6.7% | 1 | 2.2% | 0.63 |
| **Total** | 375 | 44 | 11.7% | 9 | 2.4% | <0.001 |
| **Fisher's Exact test of homogeneity, p-value<sup>c</sup>** |  | p = 0.01 |  | p = 0.08 |  |  |
| After the second dose of vaccine |  |  |  |  |  |  |
| **Pfizer-BioNTech<sup>a</sup> × 2** | 176 | 24 | 13.6% | 1 | 0.6% | <0.001 |
| **Moderna<sup>a</sup> × 2** | 35 | 5 | 14.3% | 0 | 0% | 0.06 |
| **Sputnik V<sup>b</sup> × 2** | 38 | 6 | 15.8% | 3 | 7.9% | 0.51 |
| **Astra-Zeneca<sup>b</sup> × 2** | 25 | 1 | 4.0% | 1 | 4.0% | 1 |
| **Total** | 274 | 36 | 13.1% | 5 | 1.8% | <0.001 |
| **Fisher's Exact test of homogeneity, p-value<sup>c</sup>** |  | p = 0.54 |  | p = 0.02 |  |  |

Note: The most common 4 brands were included. All participants who received the first two doses of the same vaccine were included in the analysis of post-vaccination changes after the first dose of vaccine. <sup>a</sup>mRNA type vaccines; <sup>b</sup>adenovirus vector vaccines. <sup>c</sup>Fisher's test of homogeneity was used to compare the proportions of improvement or worsening between all groups. <sup>d</sup>Binomial tests comparing the % improvement and % worsening with two-sided p-values reported.

**Table 2.** Post-vaccination changes in pre-existing phantosmia by vaccine brands.

| After the first dose of vaccine |  |  |  |  |  |  |
| --- | --- | --- | --- | --- | --- | --- |
| Brands | Vaccinated participants | Improved phantosmia |  | Worsened phantosmia |  | Improved vs worsened<sup>d</sup> |
|  | N | N | Proportion | N | Proportion | P |
| **Pfizer-BioNTech<sup>a</sup>** | 152 | 26 | 17.1% | 4 | 2.6% | <0.001 |
| **Moderna<sup>a</sup>** | 32 | 11 | 34.4% | 3 | 9.4% | 0.06 |
| **Astra-Zeneca<sup>b</sup>** | 30 | 6 | 20.0% | 0 | 0% | 0.03 |
| **Sputnik V<sup>b</sup>** | 23 | 6 | 26.1% | 2 | 8.7% | 0.29 |
| **Total** | 237 | 49 | 20.7% | 9 | 3.8% | <0.001 |
| **Fisher's Exact test of homogeneity, p-value<sup>c</sup>** |  | p = 0.15 |  | p = 0.09 |  |  |
| After the second dose of vaccine |  |  |  |  |  |  |
| **Pfizer-BioNTech<sup>a</sup> × 2** | 119 | 21 | 17.6% | 6 | 5.0% | 0.006 |
| **Moderna<sup>a</sup> × 2** | 22 | 7 | 31.8% | 1 | 4.5% | 0.07 |
| **Astra-Zeneca<sup>b</sup> × 2** | 18 | 1 | 5.6% | 0 | 0% | 1 |
| **Sputnik V<sup>b</sup> × 2** | 14 | 5 | 35.7% | 0 | 0% | 0.06 |
| **Total** | 173 | 34 | 19.7% | 9 | 5.2% | <0.001 |
| **Fisher's exact test of homogeneity, p-value<sup>c</sup>** |  | p = 0.07 |  | p = 1 |  |  |

Note: The most common 4 brands were included. All participants who received the first two doses of the same vaccine were included in the analysis of post-vaccination changes after the first dose of vaccine. <sup>a</sup>mRNA type vaccines; <sup>b</sup>adenovirus vector vaccines. <sup>c</sup>Fisher's test of homogeneity was used to compare the proportions of improvement or worsening between all groups. <sup>d</sup>Binomial tests comparing the % improvement and % worsening with two-sided p-values reported.

**Table 3.**
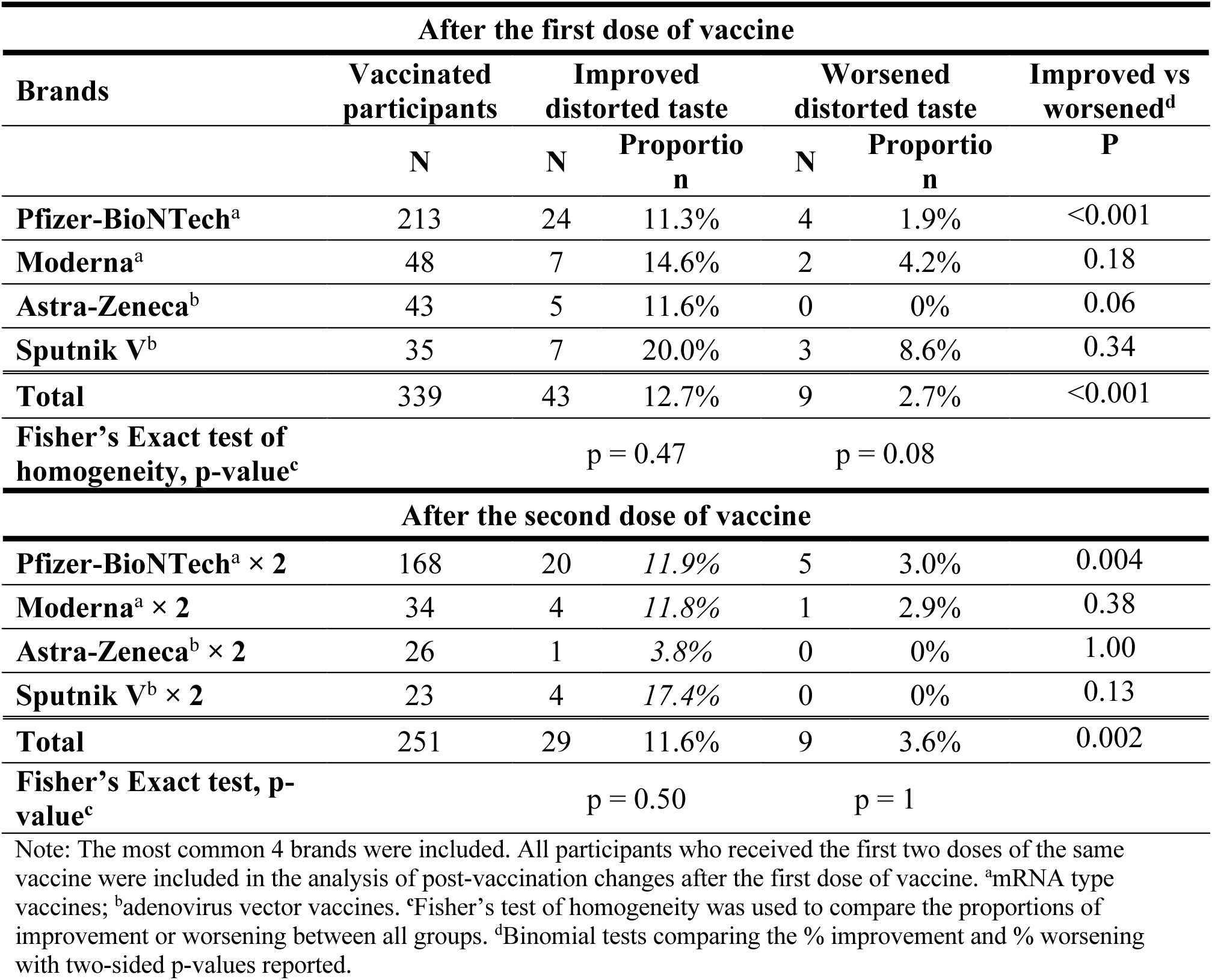
Post-vaccination changes in pre-existing distorted taste by vaccine brands.

### Side effects of vaccination

Across over 7,000 self-reported instances of post-vaccination side effects, the six most reported symptoms were fatigue (34.6% after first dose), headache (32.6%), fever (32.0%), muscle pain (29.0%), chills (25.2%), and joint pain (22.1%). Other reported symptoms included menstrual changes (10% among cis women after the first dose), brain fog (6%), and sleep disturbance (5%) (**Supplementary Figure 5**; **Supplementary Table 15**).

Side-effect incidence generally decreased with successive doses, with patterns varying by vaccine types. Among participants receiving the same vaccine type across the first three doses (*i.e.*, two doses and one booster), incidence rates for common side effects did not differ across doses in the mRNA vaccine group (n = 320; p > 0.05). In contrast, significant reductions were observed for fever and chills in the adenovirus vector vaccine group (n = 80) and headache in the inactivated virus vaccine group (n = 51) (p < 0.05) (**Supplementary Figure 6; Supplementary Tables 16and 17**).

Finally, the total number of reported side effects differed by chemosensory outcome. Participants reporting worsening of smell or taste experienced a significantly higher number of side effects compared with those reporting improvement and no change, for both olfactory and gustatory modalities (**Supplementary Figure 7**; **Supplementary Table 18**).

## DISCUSSION

In this global retrospective survey study, we examined post-vaccination changes in chemosensory perception among individuals with and without COVID-19-related chemosensory impairment. More than 90% of participants with pre-existing COVID-19-related smell or taste impairment reported no changes in their general chemosensory function (*i.e.*, the ability to smell and taste) following vaccination. Among participants with qualitative chemosensory distortions, a minority reported improvement, with over 20% reporting improvement in phantosmia, 11–18% in parosmia, and 12-21% in distorted taste, depending on vaccine dose. Worsening of chemosensory symptoms was uncommon across all modalities. Overall, these findings suggest that COVID-19 vaccination is unlikely to adversely affect chemosensory function for most individuals and are consistent with prior reports describing low-frequency post-vaccination chemosensory changes [27–29,26].

These results help reconcile differences across the existing literature on post-vaccination chemosensory outcomes. Large electronic medical record-based analyses have reported very low incidence rates of post-vaccination smell or taste disturbance [25], likely reflecting limited capture of chemosensory symptoms, particularly qualitative distortions, in routine clinical coding. In contrast, cohort studies focused on individuals with PCS have described modest changes in chemosensory symptoms following vaccination, including both improvement and worsening [26–29]. By incorporating detailed self-reported phenotyping of multiple chemosensory modalities, including parosmia and phantosmia, our study provides greater sensitivity to qualitative symptom changes that are often underrepresented in electronic health records. Differences in study design and outcome ascertainment, therefore, likely account for variability across reports, supporting the view that these findings are complementary rather than conflicting.

To further contextualize these chemosensory findings, we examined post-vaccination side effects across vaccine types and assessed whether overall side-effect burden differed by chemosensory outcome. The frequencies of reported side effects generally decreased with the vaccine doses for most vaccine types, whereas side effects following mRNA vaccines were most pronounced after the second dose. In addition to commonly reported symptoms, menstrual cycle changes were reported by more than 8% of female participants after each of the first three vaccine doses. Although prior work has suggested that menstrual cycle changes may be related to the timing of vaccination [34], our findings are observational and should be interpreted with caution. Brain fog, a symptom commonly reported in PCS, was consistently reported by 4-6% of participants across vaccine doses. While previous population-level studies suggest that vaccination may reduce the odds of brain fog as a PCS symptom [35], further longitudinal and mechanistic investigation is needed to clarify whether and how vaccination relates to cognitive symptoms [36].

From a public health perspective, these findings may help contextualize concerns about potential chemosensory side effects of COVID-19 vaccination, particularly among individuals with persistent post-COVID symptoms. The observation that chemosensory function remains unchanged for most individuals, with improvement occurring in a subset of those with qualitative chemosensory distortions, highlights the heterogeneity of post-COVID sensory trajectories. Although the pathophysiological mechanisms underlying vaccine-associated changes in chemosensory perception remain unclear [37], clear communication of these patterns may support informed decision-making and help address vaccine hesitancy in populations affected by post-COVID chemosensory dysfunction, without implying vaccination as a therapeutic intervention.

Several limitations should be considered when interpreting these findings. Recruitment through social media and recontacting participants from prior chemosensory studies may have introduced selection bias, potentially overrepresenting individuals with more persistent symptoms or greater motivation to report outcomes. Self-selection bias is also possible, which may limit the generalizability to the broader vaccinated population. All outcomes were self-reported, raising the possibility of recall bias and misclassification. Although self-report methods have been widely used to capture chemosensory outcomes in COVID-19 research [15–17], they lack the precision of standardized psychophysical assessments [24]. Additionally, COVID-19-related chemosensory dysfunction often improves gradually over time, making it difficult to distinguish vaccine-related changes from natural recovery. While we observed no association between time since vaccination and reported chemosensory changes, the absence of longitudinal tracking and objective baseline measures limits causal inference. The survey period spanned multiple SARS-CoV-2 variants and infection waves, which may differ in their chemosensory sequelae. Finally, cross-country differences in healthcare access, vaccine availability, and cultural norms may have influenced both exposure and symptom reporting. Collectively, these limitations indicate that the findings are exploratory and should be interpreted with caution.

## CONCLUSIONS

This global survey provides evidence that COVID-19 vaccination is associated with low-frequency changes in chemosensory perception among individuals with prior COVID-19-related chemosensory dysfunction, particularly for qualitative symptoms such as parosmia, phantosmia, and distorted taste. Most participants reported no change in their general sense of smell or taste following vaccination, and worsening of chemosensory symptoms was uncommon. Although the observational design precludes causal inference, these findings suggest that vaccination is unlikely to be associated with adverse chemosensory outcomes for most individuals and may coincide with improvement in a subset experiencing qualitative distortions. Further longitudinal studies with objective assessments (e.g., Sniffn’ Sticks [38] or taste strips [39]) are needed to confirm these observations and to better inform public health communication around vaccination and post-COVID sensory symptoms.

## Data Availability

All data produced in the present study are available upon reasonable request to the authors.

## ACKNOWLEDGEMENTS

LDH is supported by an Australian Research Council Early Discovery Researcher Award (DE240100014). We would like to thank Lisa Arvieu for assisting in translating the data collected from the French survey into English and Alyssa Bakke for emailing the survey to participants of previous GCCR studies.

## AUTHORSHIP CONTRIBUTION

SK1, CMC, SK2, EC, MDG, JC, VP, VVV and LDH conceived the study. SK1, CMC, SK2, EC, MDG, JC, VP, KO, VVV and LDH designed the original survey. SK1, CMC, SK2, EC, SP, MDG, VH, SC, VP, JC, GC, KO, VVV and LDH translated and implemented the survey. SK1 and LDH set up the online survey. SK1, CMC, MAK, TKL, SK2, IGK, JY, MDG, VVV and LDH analyzed data. SK1, CMC, MAK, TKL, SK2, IGK, JY, MDG, VP, VVV and LDH interpreted data. SK1, CMC, SK2, EC, MDG, VP, JC, VVV and LDH administrated project. SK1, CMC, MAK, TKL, SK2, IGK, EC, MDG, VVV and LDH wrote the original manuscript. All authors reviewed and approved the final version of the manuscript. SK1, Sachiko Koyama; SK2, Svetlana Kopishinskaia.

## DATA AVAILABILITY

The data underlying this article will be shared on reasonable request to the corresponding author.

