## Supplementary Materials for "Post-vaccination chemosensory outcomes in COVID-19-associated dysfunction"

### Supplementary Note

Brand of COVID-19 vaccines grouped by type:
1) **mRNA vaccines**: Pfizer–BioNTech and Moderna.
2) **Adenovirus vector vaccines**: Oxford–AstraZeneca COVID-19 vaccine, the Sputnik V COVID-19 vaccine, Sputnik Light, Covishield (Serum Ins., India), Convidecia, and the Janssen COVID-19 vaccine.
3**) Inactivated virus vaccines**: Chinese CoronaVac and the Sinopharm BIBP and WIBP vaccines; the Indian Covaxin; the Russian CoviVac; the Kazakh vaccine QazVac; the Iranian COVIran Barekat, Valneva COVID-19 vaccine.
4) **Recombinant protein vaccines**:

i) Subunit vaccines: the peptide vaccine EpiVacCorona, ZF2001, MVC-COV1901, Corbevax, the Sanofi–GSK vaccine, and Soberana 02 (a conjugate vaccine). Bimervax was approved for use as a booster vaccine in the European Union in March 2023.

ii) Virus-like particle vaccines: Novavax COVID-19 vaccine.

### Supplementary Tables

**Supplementary Table 1. Number of vaccinated participants from each country.**

| Country | Number of Participants per country listed |
|---|---|
| Russian Federation | 245 (18%) |
| France | 225 (17%) |
| Iran | 133 (10% |
| Italy | 125 (9%) |
| United States of America | 112 (8.45%) |
| Spain | 94 (7.09%) |
| Australia | 92 (6.94%) |
| Germany | 85 (6.4%) |
| United Kingdom of Great Britain and Northern Ireland | 36 (2.7%) |
| Canada, Switzerland | 24 (1.8%)* |
| Japan | 21 (1.58%) |
| Kazakhstan | 13 (0.98%) |
| Austria | 9 0.68%) |
| Belgium, China, Mexico | 8 (0.6%)* |
| Taiwan | 7 (0.5%) |
| Peru | 5 (0.37%) |
| Argentina, Bulgaria, Chile, Colombia, Hungary, Netherlands, South Africa | 3 (0.2%)* |
| Brazil, Kyrgyzstan, Luxembourg, Mauritius, Sweden, Ukraine | 2 (0.15%) |
| Afghanistan, Algeria, Belarus, Brunei Darussalam, Denmark, Ecuador, Hong Kong, India, Indonesia, Ireland, Israel, Kenya, Kuwait, Lebanon, Morocco, New Zealand, Nigeria, Republic of Moldova, Timor-Leste | 1 (0.07%)* |
| **TOTAL** | 1326 |

*per each of the referred countries

We note that 1326 of 1352 participants reported their country of residence.

**Supplementary Table 2. Participants' responses to questions about their sense of smell after each dose of vaccine.**

| Answer options | Dose 1 | | Dose 2 | | Booster 1 | | Booster 2 | |
|---|---|---|---|---|---|---|---|---|
| | N | % | N | % | N | % | N | % |
| **HOW WAS YOUR SMELL BEFORE VACCINATION?** | | | | | | | | |
| Normal | 618 | 46% | 658 | 54% | 485 | 57% | 136 | 57% |
| Affected by COVID-19 / fully recovered | 226 | 17% | 151 | 12% | 111 | 13% | 31 | 13% |
| Affected by COVID-19 / partially recovered | 259 | 19% | 231 | 19% | 141 | 17% | 38 | 16% |
| Affected by COVID-19 / not recovered | 117 | 9% | 103 | 8% | 56 | 7% | 16 | 7% |
| Loss of smell due to COVID-19 | 100 | 7% | 61 | 5% | 41 | 5% | 10 | 4% |
| Other | 32 | 2% | 23 | 2% | 19 | 2% | 7 | 3% |
| Total | 1352 | | 1227 | | 853 | | 238 | |
| **DID THE VACCINATION AFFECT YOUR SENSE OF SMELL?** | | | | | | | | |
| Yes, sense worsened | 35 | 3% | 28 | 2% | 15 | 2% | 6 | 3% |
| Yes, sense improved | 44 | 3% | 32 | 3% | 30 | 4% | 6 | 3% |
| No | 1183 | 94% | 1101 | 95% | 751 | 94% | 207 | 94% |
| Total | 1262 | | 1161 | | 796 | | 219 | |
| **IF VACCINATION AFFECTED YOUR SENSE OF SMELL, WHEN DID THE CHANGE START?** | | | | | | | | |
| Immediately after vaccination | 8 | 4% | 4 | 2% | 6 | 5% | 2 | 7% |
| Within 1 day of vaccination | 18 | 9% | 13 | 7% | 3 | 2% | 1 | 3% |
| After 1 day of vaccination | 49 | 25% | 39 | 22% | 31 | 25% | 9 | 30% |
| The vaccination did not affect my sense of smell | 121 | 62% | 121 | 68% | 82 | 67% | 18 | 60% |
| Total | 196 | | 177 | | 122 | | 30 | |
| **IF VACCINATION AFFECTED YOUR SENSE OF SMELL, HOW LONG DID THE CHANGE LAST?** | | | | | | | | |
| <1 day | 6 | 3% | 4 | 2% | 4 | 3% | 2 | 7% |
| 1 day to 1 week | 11 | 6% | 5 | 3% | 2 | 2% | 0 | 0% |
| 1 week to 1 month | 11 | 6% | 5 | 3% | 2 | 2% | 3 | 10% |
| >1 month | 43 | 22% | 46 | 26% | 31 | 26% | 8 | 27% |
| No effect | 124 | 64% | 118 | 66% | 81 | 68% | 17 | 57% |
| Total | 195 | | 178 | | 120 | | 30 | |
| **DID YOU EXPERIENCE PAROSMIA AFTER YOUR VACCINATION?** | | | | | | | | |
| Had it before vaccination / worsened after vaccination | 11 | 1% | 7 | 1% | 5 | 1% | 1 | 0% |
| Had it before vaccination / did not change after vaccination | 366 | 29% | 301 | 26% | 182 | 22% | 44 | 19% |
| Had it before vaccination / improved after vaccination | 47 | 4% | 48 | 4% | 27 | 3% | 10 | 4% |
| Did not have it before vaccination / started after vaccination | 11 | 1% | 10 | 1% | 8 | 1% | 3 | 1% |

| | | | | | | | | |
|---|---|---|---|---|---|---|---|---|
| Did not have it before or after vaccination | 845 | 66% | 803 | 69% | 602 | 73% | 179 | 76% |
| Total | 1280 | | 1169 | | 824 | | 237 | |
| **DID YOU EXPERIENCE PHANTOSMIA AFTER YOUR VACCINATION?** | | | | | | | | |
| Had it before vaccination / worsened after vaccination | 9 | 1% | 7 | 1% | 3 | 0% | 1 | 0% |
| Had it before vaccination / did not change after vaccination | 203 | 16% | 162 | 14% | 98 | 12% | 26 | 11% |
| Had it before vaccination / improved after vaccination | 52 | 4% | 49 | 4% | 36 | 4% | 11 | 5% |
| Did not have it before vaccination / started after vaccination | 23 | 2% | 22 | 2% | 18 | 2% | 5 | 2% |
| Did not have it before or after vaccination | 979 | 77% | 921 | 79% | 661 | 81% | 190 | 82% |
| Total | 1266 | | 1161 | | 816 | | 233 | |

**Supplementary Table 3. Participants' responses to questions about their sense of taste and chemesthesis after each dose of vaccine.**

| Answer options | Dose 1 | | Dose 2 | | Booster 1 | | Booster 2 | |
|---|---|---|---|---|---|---|---|---|
| | N | % | N | % | N | % | N | % |
| **HOW WAS YOUR SENSE OF TASTE BEFORE VACCINATION?** | | | | | | | | |
| Normal | 712 | 54% | 719 | 59% | 537 | 63% | 151 | 63% |
| Affected by COVID-19 / fully recovered | 255 | 19% | 194 | 16% | 123 | 14% | 34 | 14% |
| Affected by COVID-19 / partially recovered | 196 | 15% | 174 | 14% | 113 | 13% | 25 | 10% |
| Affected by COVID-19 / not recovered | 66 | 5% | 59 | 5% | 34 | 4% | 10 | 4% |
| Loss of smell due to COVID-19 | 79 | 6% | 51 | 4% | 34 | 4% | 8 | 3% |
| Other | 22 | 2% | 20 | 2% | 12 | 1% | 11 | 5% |
| Total | 1330 | | 1217 | | 853 | | 239 | |
| **DID THE VACCINATION AFFECT YOUR SENSE OF TASTE?** | | | | | | | | |
| Yes, sense worsened | 22 | 2% | 21 | 2% | 6 | 1% | 4 | 2% |
| Yes, sense improved | 31 | 3% | 19 | 2% | 20 | 3% | 6 | 3% |
| No | 1189 | 95% | 1101 | 96% | 766 | 96% | 209 | 95% |
| Total | 1242 | | 1141 | | 792 | | 219 | |
| **If VACCINATION AFFECTED YOUR SENSE OF TASTE, WHEN DID THE CHANGE START?** | | | | | | | | |
| Immediately after vaccination | 1 | 1% | 1 | 1% | 2 | 2% | 1 | 4% |
| Within 1 day of vaccination | 13 | 8% | 11 | 7% | 4 | 4% | 1 | 4% |
| After 1 day of vaccination | 36 | 22% | 27 | 18% | 21 | 19% | 8 | 33% |
| The vaccination did not affect my sense of smell | 116 | 70% | 115 | 75% | 81 | 75% | 14 | 58% |
| Total | 166 | | 154 | | 108 | | 24 | |
| **IF VACCINATION AFFECTED YOUR SENSE OF TASTE, HOW LONG DID THE CHANGE LAST?** | | | | | | | | |
| <1 day | 2 | 1% | 2 | 1% | 3 | 3% | 1 | 4% |
| 1 day to 1 week | 9 | 5% | 5 | 3% | 2 | 2% | 1 | 4% |
| 1 week to 1 month | 7 | 4% | 5 | 3% | 4 | 4% | 3 | 13% |
| >1 month | 35 | 21% | 30 | 19% | 21 | 19% | 6 | 26% |
| No effect | 114 | 68% | 112 | 73% | 80 | 73% | 12 | 52% |
| Total | 167 | | 154 | | 110 | | 23 | |
| **ABOUT DISTORTED TASTE?** | | | | | | | | |
| Had it before vaccination / worsened after vaccination | 10 | 1% | 6 | 1% | 2 | 0% | 1 | 0% |
| Had it before vaccination / did not change after vaccination | 325 | 26% | 282 | 25% | 170 | 22% | 37 | 16% |
| Had it before vaccination / improved after vaccination | 45 | 4% | 38 | 3% | 29 | 4% | 10 | 4% |
| Did not have it before vaccination / started after vaccination | 16 | 1% | 12 | 1% | 12 | 2% | 2 | 1% |

| | | | | | | | | |
|---|---|---|---|---|---|---|---|---|
| Did not have it before or after vaccination | 836 | 68% | 792 | 70% | 574 | 73% | 177 | 78% |
| Total | 1232 | | 1130 | | 787 | | 227 | |
| **DID THE VACCINATION AFFECT YOUR SENSE OF CHEMESTHESIS?** | | | | | | | | |
| Yes, sense worsened | 18 | 2% | 18 | 2% | 12 | 2% | 3 | 1% |
| Yes, sense improved | 9 | 1% | 8 | 1% | 8 | 1% | 3 | 1% |
| No | 1159 | 98% | 1108 | 98% | 760 | 97% | 215 | 97% |
| Total | 1186 | | 1134 | | 780 | | 221 | |

**Supplementary Table 4. Proportion of chemosensory impairments prior to the first dose of vaccine by countries.**

| Country | Impaired sense? | General smell | | General taste | | Parosmia | | Phantosmia | | Distorted taste | |
|---|---|---|---|---|---|---|---|---|---|---|---|
| | | n | % | n | % | n | % | n | % | n | % |
| **Russia** | NO | 190 | 78.8% | 215 | 90.3% | 183 | 77.5% | 209 | 88.9% | 203 | 84.9% |
| | YES | 51 | 21.2% | 23 | 9.7% | 53 | 22.5% | 26 | 11.1% | 36 | 15.1% |
| **France** | NO | 111 | 50.5% | 141 | 65.9% | 103 | 49.3% | 139 | 66.5% | 105 | 50.5% |
| | YES | 109 | 49.5% | 73 | 34.1% | 106 | 50.7% | 70 | 33.5% | 103 | 49.5% |
| **Iran** | NO | 113 | 91.1% | 125 | 97.7% | 108 | 95.6% | 105 | 97.2% | 79 | 95.2% |
| | YES | 11 | 8.9% | 3 | 2.3% | 5 | 4.4% | 3 | 2.8% | 4 | 4.8% |
| **Italy** | NO | 73 | 58.9% | 89 | 72.4% | 86 | 71.1% | 99 | 81.8% | 86 | 74.1% |
| | YES | 51 | 41.1% | 34 | 27.6% | 35 | 28.9% | 22 | 18.2% | 30 | 25.9% |
| **USA** | NO | 44 | 40.0% | 53 | 48.2% | 61 | 56.0% | 68 | 63.0% | 59 | 54.1% |
| | YES | 66 | 60.0% | 57 | 51.8% | 48 | 44.0% | 40 | 37.0% | 50 | 45.9% |
| **Spain** | NO | 41 | 45.6% | 48 | 52.7% | 43 | 48.9% | 64 | 71.9% | 46 | 52.9% |
| | YES | 49 | 54.4% | 43 | 47.3% | 45 | 51.1% | 25 | 28.1% | 41 | 47.1% |
| **Australia** | NO | 91 | 98.9% | 91 | 98.9% | 89 | 98.9% | 87 | 97.8% | 87 | 98.9% |
| | YES | 1 | 1.1% | 1 | 1.1% | 1 | 1.1% | 2 | 2.3% | 1 | 1.1% |
| **Germany** | NO | 42 | 50.6% | 51 | 62.2% | 44 | 53.0% | 56 | 70.0% | 50 | 61.7% |
| | YES | 41 | 49.4% | 31 | 37.8% | 39 | 47.0% | 24 | 30.0% | 31 | 38.3% |
| **UK** | NO | 6 | 17.1% | 11 | 30.6% | 12 | 34.3% | 20 | 57.1% | 11 | 31.4% |
| | YES | 29 | 82.9% | 25 | 69.4% | 23 | 65.7% | 15 | 42.9% | 24 | 68.6% |
| **Canada** | NO | 5 | 21.7% | 5 | 23.8% | 8 | 34.8% | 10 | 43.5% | 8 | 36.4% |
| | YES | 18 | 78.3% | 16 | 76.2% | 15 | 65.2% | 13 | 56.5% | 14 | 63.6% |
| **Switzerland** | NO | 12 | 50.0% | 15 | 65.2% | 16 | 69.6% | 20 | 83.3% | 17 | 73.9% |
| | YES | 12 | 50.0% | 8 | 34.8% | 7 | 30.4% | 4 | 16.7% | 6 | 26.1% |
| **Japan** | NO | 21 | 100.0% | 21 | 100.0% | 20 | 95.2% | 21 | 100.0% | 21 | 100.0% |
| | YES | | | | | 1 | 4.8% | | | | |
| **Kazakhstan** | NO | 8 | 66.7% | 12 | 100.0% | 7 | 53.8% | 8 | 80.0% | 10 | 76.9% |
| | YES | 4 | 33.3% | | | 6 | 46.2% | 2 | 20.0% | 3 | 23.1% |
| **Total (impaired)** | | **1199 (442)** | | **1191 (314)** | | **1164 (384)** | | **1152 (246)** | | **1125 (343)** | |

Only countries with 10 or more participants are included.

**Supplementary Table 5. Combinations of the first three doses by vaccine brands in different countries.**

| Country | P+P+P | P+P+M | SV+SV+SL | M+M+M | S+S+S | A+A+P | A+A+M | SV+SV+SV | M+M+P | S+S+A | A+P+P | A+A+A | Other combinations |
|---|---|---|---|---|---|---|---|---|---|---|---|---|---|
| France | 72 (63.7%) | 16 (14.2%) | 0 | 4 (3.5%) | 0 | 1 (0.9%) | 1 (0.9%) | 0 | 5 (4.4%) | 0 | 7 (6.2%) | 0 | 7 (6.2%) |
| Russia | 0 | 0 | 58 (55.8%) | 0 | 0 | 0 | 0 | 30 (28.8%) | 0 | 0 | 0 | 0 | 16 (15.4%) |
| Italy | 43 (43.0%) | 29 (29.0%) | 0 | 5 (5.0%) | 0 | 2 (2.0%) | 8 (8.0%) | 0 | 4 (4.0%) | 0 | 2 (2.0%) | 0 | 7 (7.0%) |
| Iran | 2 (2.4%) | 0 | 0 | 0 | 35 (41.2%) | 0 | 0 | 0 | 0 | 16 (18.8%) | 0 | 7 (8.2%) | 25 (29.4%) |
| Australia | 41 (53.2%) | 11 (14.3%) | 0 | 1 (1.3%) | 0 | 14 (18.2%) | 7 (9.1%) | 0 | 0 | 0 | 0 | 1 (1.3%) | 2 (2.6%) |
| USA | 37 (50.7%) | 6 (8.2%) | 0 | 22 (30.1%) | 0 | 0 | 1 (1.4%) | 0 | 2 (2.7%) | 0 | 0 | 0 | 5 (6.8%) |
| Germany | 19 (32.8%) | 13 (22.4%) | 0 | 2 (3.4%) | 0 | 4 (6.9 | 4 (6.9%) | 0 | 2 (3.4%) | 0 | 3 (5.2%) | 0 | 11 (19.0%) |
| Spain | 13 (31.7%) | 19 (46.3%) | 0 | 3 (7.3%) | 0 | 0 | 2 (4.9%) | 0 | 1 (2.4%) | 0 | 0 | 0 | 3 (7.3%) |
| UK | 8 (27.6%) | 1 (3.4%) | 0 | 0 | 0 | 9 (31.0%) | 4 (13.8%) | 0 | 3 (10.3%) | 0 | 1 (3.4%) | 0 | 3 (10.3%) |
| Canada | 7 (38.9%) | 4 (22.2%) | 0 | 1 (5.6%) | 0 | 1 (5.6%) | 0 | 0 | 0 | 0 | 1 (5.6%) | 0 | 4 (22.2%) |
| Japan | 13 (81.3%) | 2 (12.5%) | 0 | 0 | 0 | 0 | 0 | 0 | 1 (6.3%) | 0 | 0 | 0 | 0 |
| Switzerland | 3 (27.3%) | 1 (9.1%) | 0 | 5 (45.5%) | 0 | 0 | 0 | 0 | 2 (18.2%) | 0 | 0 | 0 | 0 |
| Others* | 17 (21.8%) | 6 (7.7%) | 1 (1.3%) | 1 (1.3%) | 1 (1.3%) | 5 (6.4%) | 4 (5.1%) | 0 | 3 (3.8%) | 1 (1.3%) | 1 (1.3%) | 2 (2.6%) | 37 (46.8%) |
| **Total** | 275 (34.2%) | 108 (13.4%) | 59 (7.3%) | 44 (5.5%) | 36 (4.5%) | 36 (4.5%) | 31 (3.9%) | 30 (3.7%) | 23 (2.9%) | 17 (2.1%) | 15 (1.9%) | 10 (1.2%) | 120 (14.9%) |

*Others represent countries with the sum of vaccinated participants < 10.

P, Pfizer-BioNTech; M, Moderna; SV, Sputnik V; SL, Sputnik Light; S, Sinopharm; A, Astra-Zeneca;

**Supplementary Table 6. Comparison of changes in the senses of smell and taste after each dose of vaccine in participants with and without pre-existing sensory impairment due to COVID-19.**

| Vaccine dose | Improvement in individuals with pre-existing impairment | | | Improvement in individuals without pre-existing impairment | | | Fisher's exact test (p-value) |
|---|---|---|---|---|---|---|---|
| | Total N | N | Proportion | Total N | N | Proportion | |
| **General sense of smell** | | | | | | | |
| **Dose 1** | 452 | 33 | 7.3% | 785 | 8 | 1.0% | $6.092 \times 10^{-9}$ |
| **Dose 2** | 373 | 20 | 5.4% | 756 | 9 | 1.2% | $7.337 \times 10^{-5}$ |
| **Dose 3** | 223 | 17 | 7.6% | 549 | 9 | 1.6% | $9.266 \times 10^{-5}$ |
| **Dose 4** | 60 | 3 | 5.0% | 150 | 2 | 1.3% | 0.1421 |
| **General sense of taste** | | | | | | | |
| **Dose 1** | 312 | 23 | 7.4% | 900 | 6 | 0.7% | $1.447 \times 10^{-9}$ |
| **Dose 2** | 262 | 14 | 5.3% | 842 | 3 | 0.4% | $4.557 \times 10^{-7}$ |
| **Dose 3** | 166 | 12 | 7.2% | 605 | 6 | 1.0% | $3.853 \times 10^{-5}$ |
| **Dose 4** | 40 | 1 | 2.5% | 167 | 4 | 2.4% | 1 |

| Vaccine dose | Worsening in individuals with pre-existing impairment | | | Worsening in individuals without pre-existing impairment | | | Fisher's exact test (p-value) |
|---|---|---|---|---|---|---|---|
| | Total N | N | Proportion | Total N | N | Proportion | |
| **General sense of smell** | | | | | | | |
| **Dose 1** | 452 | 13 | 2.9% | 785 | 19 | 2.4% | 0.7104 |
| **Dose 2** | 373 | 15 | 4.0% | 756 | 9 | 1.2% | 0.0034 |
| **Dose 3** | 223 | 9 | 4.0% | 549 | 4 | 0.7% | 0.0028 |
| **Dose 4** | 60 | 4 | 6.7% | 150 | 1 | 0.7% | 0.0242 |
| **General sense of taste** | | | | | | | |
| **Dose 1** | 312 | 13 | 4.2% | 900 | 7 | 0.8% | 0.0002 |
| **Dose 2** | 262 | 13 | 5.0% | 842 | 5 | 0.6% | $1.567 \times 10^{-5}$ |
| **Dose 3** | 166 | 4 | 2.4% | 605 | 1 | 0.2% | 0.0087 |
| **Dose 4** | 40 | 2 | 5.0% | 167 | 1 | 0.6% | 0.0962 |

**Supplementary Table 7. Durations between survey date and vaccination by post-vaccination changes in chemosensory perception in participants with pre-existing distortion**

| Time between the survey and Dose 1 by general smell changes (after dose 1) | |
|---|---|
| **Change in smell** | **Mean (standard deviation) in month** |
| Worsened (n = 13) | 20.1 (5.60) |
| Improved (n = 33) | 20.2 (4.37) |
| No change (n = 402) | 20.4 (5.32) |
| ANOVA results: F (1, 446) = 0.135, p = 0.714 | |

| Time between the survey and Dose 1 by general taste changes (after dose 1) | |
|---|---|
| **Change in taste** | **Mean (standard deviation) in month** |
| Worsened (n = 13) | 18.5 (4.93) |
| Improved (n = 23) | 20.9 (4.35) |
| No change (n = 273) | 20.9 (5.32) |
| ANOVA results: F (1, 307) = 1.78, p = 0.183 | |

| Time between the survey and Dose 1 by parosmia changes (after dose 1) | |
|---|---|
| **Change in parosmia** | **Mean (standard deviation) in month** |
| Worsened (n = 11) | 19.8. (9.37) |
| Improved (n = 47) | 19.2 (4.84) |
| No change (n = 362) | 20.1 (5.32) |
| ANOVA results: F (1, 418) = 0.813, p = 0.368 | |

| Time between the survey and Dose 1 by phantosmia changes (after dose 1) | |
|---|---|
| **Change in phantosmia** | **Mean (standard deviation) in month** |
| Worsened (n = 9) | 18.4 (7.18) |
| Improved (n = 52) | 19.6 (4.56) |
| No change (n = 201) | 21.0 (5.33) |
| ANOVA results: F (1, 260) = 0.793, p = 0.374 | |

| Time between the survey and Dose 1 by distorted taste changes (after dose 1) | |
|---|---|
| **Change in distorted taste** | **Mean (standard deviation) in month** |
| Worsened (n = 10) | 17.5 (5.50) |
| Improved (n = 45) | 20.1 (5.42) |
| No change (n = 321) | 20.6 (5.30) |
| ANOVA results: F (1, 374) = 0.146, p = 0.702 | |

**Supplementary Table 8. Changes in smell after the first and the second dose of the five most common vaccine brands.**

| Brands | Vaccinated participants | Improved smell | | Worsened smell | | Improved vs worsened[e] |
|---|---|---|---|---|---|---|
| **After the first dose of vaccine** | | | | | | |
| | N | N | Proportion | N | Proportion | P |
| **Pfizer-BioNTech[a]** | 598 | 18 | 3.0% | 8 | 1.3% | 0.08 |
| **Sputnik V[b]** | 246 | 2 | 0.8% | 11 | 4.5% | 0.02 |
| **Astra-Zeneca[b]** | 159 | 7 | 4.4% | 7 | 4.4% | 1 |
| **Moderna[a]** | 125 | 12 | 9.6% | 4 | 3.2% | 0.08 |
| **Sinopharm[c]** | 108 | 2 | 1.9% | 2 | 1.9% | 1 |
| **Total** | 1236 | 41 | 3.3% | 32 | 2.6% | 0.35 |
| **Fisher's Exact test of homogeneity, p-value [d]** | | p < 0.001 | | p = 0.03 | | |
| **After the second dose of vaccine** | | | | | | |
| **Pfizer-BioNTech[a] × 2** | 529 | 14 | 2.6% | 4 | 0.8% | 0.03 |
| **Sputnik V[b] × 2** | 194 | 3 | 1.5% | 9 | 4.6% | 0.15 |
| **Astra-Zeneca[b] × 2** | 108 | 3 | 2.8% | 3 | 2.8% | 1 |
| **Sinopharm[c] × 2** | 102 | 1 | 1.0% | 2 | 2.0% | 1 |
| **Moderna[a] × 2** | 99 | 7 | 7.1% | 5 | 5.1% | 0.77 |
| **Total** | 1032 | 28 | 2.7% | 23 | 2.2% | 0.58 |
| **Fisher's Exact test of homogeneity, p-value[d]** | | p = 0.09 | | p = 0.002 | | |

Note: Brands with ≥95 respondents were included. All participants who received the first two doses of the same vaccine were included in the analysis of post-vaccination changes after the first dose of vaccine. [a]mRNA type vaccines; [b]adenovirus vector vaccines; [c]inactivated virus vaccines. [d]Fisher's test of homogeneity was used to compare the proportions of improvement or worsening between all groups. [e]Binomial tests comparing the % improvement and % worsening with two-sided p-values reported.

**Supplementary Table 9a**. Changes in smell after the first dose of the five most common vaccine brands **by countries**

| Country | N | Pfizer-BioNTech | | | | | Sputnik V | | | | | AstraZeneca | | | | | Moderna | | | | | Sinopharm | | | | | Total | | | | |
|---|---|---|---|---|---|---|---|---|---|---|---|---|---|---|---|---|---|---|---|---|---|---|---|---|---|---|---|---|---|---|---|
| | | TV | SW | SW% | SI | SI% | TV | SW | SW% | SI | SI% | TV | SW | SW% | SI | SI% | TV | SW | SW% | SI | SI% | TV | SW | SW% | SI | SI% | TV | SW | SW% | SI | SI% |
| **Russia** | **245** | 1 | 0 | 0% | 0 | 0% | 211 | 9 | 4% | 2 | 1% | 0 | 0 | na | 0 | na | 0 | 0 | na | 0 | na | 0 | 0 | na | 0 | na | **212** | **9** | **4%** | **2** | **1%** |
| **France** | **225** | 184 | 4 | 2% | 5 | 3% | 0 | 0 | na | 0 | na | 16 | 0 | 0% | 2 | 13% | 22 | 1 | 5% | 2 | 9% | 0 | 0 | na | 0 | na | **222** | **5** | **2%** | **9** | **4%** |
| **Iran** | **133** | 3 | 0 | 0% | 0 | 0% | 14 | 0 | 0% | 0 | 0% | 25 | 3 | 12% | 0 | 0% | 0 | 0 | na | 0 | na | 84 | 0 | 0% | 2 | 2% | **126** | **3** | **2%** | **2** | **2%** |
| **Italy** | **125** | 85 | 0 | 0% | 3 | 4% | 0 | 0 | na | 0 | na | 20 | 1 | 5% | 1 | 5% | 15 | 0 | 0% | 4 | 27% | 0 | 0 | na | 0 | na | **120** | **1** | **1%** | **8** | **7%** |
| **USA** | **112** | 58 | 1 | 2% | 3 | 5% | 0 | 0 | na | 0 | na | 1 | 0 | 0% | 0 | 0% | 34 | 1 | 3% | 3 | 9% | 0 | 0 | na | 0 | na | **93** | **2** | **2%** | **6** | **6%** |
| **Spain** | **94** | 62 | 1 | 2% | 1 | 2% | 0 | 0 | na | 0 | na | 8 | 1 | 13% | 0 | 0% | 8 | 1 | 13% | 1 | 13% | 0 | 0 | na | 0 | na | **78** | **3** | **4%** | **2** | **3%** |
| **Australia** | **92** | 61 | 0 | 0% | 0 | 0% | 0 | 0 | na | 0 | na | 25 | 0 | 0% | 0 | 0% | 2 | 0 | 0% | 0 | 0% | 1 | 0 | 0% | 0 | 0% | **89** | **0** | **0%** | **0** | **0%** |
| **Germany** | **85** | 52 | 1 | 2% | 4 | 8% | 0 | 0 | na | 0 | na | 20 | 0 | 0% | 2 | 10% | 9 | 1 | 11% | 0 | 0% | 0 | 0 | na | 0 | na | **81** | **2** | **2%** | **6** | **7%** |
| **United Kingdom** | **36** | 12 | 1 | 8% | 0 | 0% | 0 | 0 | na | 0 | na | 17 | 0 | 0% | 1 | 6% | 6 | 0 | 0% | 1 | 17% | 0 | 0 | na | 0 | na | **35** | **1** | **3%** | **2** | **6%** |
| **Canada** | **24** | 18 | 0 | 0% | 0 | 0% | 0 | 0 | na | 0 | na | 4 | 0 | 0% | 0 | 0% | 1 | 0 | 0% | 0 | 0% | 0 | 0 | na | 0 | na | **23** | **0** | **0%** | **0** | **0%** |
| **Switzerland** | **24** | 6 | 0 | 0% | 0 | 0% | 0 | 0 | na | 0 | na | 0 | 0 | na | 0 | na | 17 | 0 | 0% | 1 | 6% | 0 | 0 | na | 0 | na | **23** | **0** | **0%** | **1** | **4%** |
| **Japan** | **21** | 19 | 0 | 0% | 0 | 0% | 0 | 0 | na | 0 | na | 1 | 0 | 0% | 0 | 0% | 1 | 0 | 0% | 0 | 0% | 0 | 0 | na | 0 | na | **21** | **0** | **0%** | **0** | **0%** |
| **Kazakhstan** | **13** | 0 | 0 | na | 0 | na | 13 | 2 | 15% | 0 | 0% | 0 | 0 | na | 0 | na | 0 | 0 | na | 0 | na | 0 | 0 | na | 0 | na | **13** | **2** | **15%** | **0** | **0%** |

Countries with more than 10 participants were considered (total N = 1229, N of vaccinated with the specific vaccines = 1136); N – number of participants per country; TV – total vaccinated, N; SW, SW% - sense worsened, N, %; SI, SI% - sense improved, N, %; na – not available.

**Supplementary Table 9b**. Changes in smell after the first dose of the five most common vaccine brands **by world regions**

| Region | N | Pfizer-BioNTech | | | | | Sputnik V | | | | | AstraZeneca | | | | | Moderna | | | | | Sinopharm | | | | | Total | | | | |
|---|---|---|---|---|---|---|---|---|---|---|---|---|---|---|---|---|---|---|---|---|---|---|---|---|---|---|---|---|---|---|---|
| | | TV | SW | SW% | SI | SI% | TV | SW | SW% | SI | SI% | TV | SW | SW% | SI | SI% | TV | SW | SW% | SI | SI% | TV | SW | SW% | SI | SI% | TV | SW | SW% | SI | SI% |
| **Europe** | **870** | 421 | 7 | 2% | 14 | 3% | 212 | 9 | 4% | 2 | 1% | 87 | 2 | 2% | 7 | 8% | 82 | 3 | 4% | 9 | 11% | 2 | 0 | 0% | 0 | 0% | **804** | **21** | **3%** | **32** | **4%** |
| **Asia** | **193** | 28 | 0 | 0% | 0 | 0% | 28 | 2 | 7% | 0 | 0% | 32 | 3 | 9% | 0 | 0% | 2 | 0 | 0% | 0 | 0% | 88 | 1 | 1% | 2 | 2% | **178** | **6** | **3%** | **2** | **1%** |
| **Americas** | **161** | 80 | 1 | 1% | 3 | 4% | 1 | 0 | 0% | 0 | 0% | 10 | 1 | 10% | 0 | 0% | 38 | 1 | 3% | 3 | 8% | 3 | 1 | 33% | 0 | 0% | **132** | **4** | **3%** | **6** | **5%** |
| **Oceania** | **93** | 62 | 0 | 0% | 0 | 0% | 0 | 0 | na | 0 | na | 25 | 0 | 0% | 0 | 0% | 2 | 0 | 0% | 0 | 0% | 1 | 0 | 0% | 0 | 0% | **90** | **0** | **0%** | **0** | **0%** |
| **Africa** | **9** | 3 | 0 | 0% | 0 | 0% | 0 | 0 | na | 0 | na | 2 | 1 | 50% | 0 | 0% | 1 | 0 | 0% | 0 | 0% | 0 | 0 | na | 0 | na | **6** | **1** | **17%** | **0** | **0%** |

All participants who indicated their country were considered (total N = 1326, N of vaccinated with the specific vaccines = 1210)

**Supplementary Table 9c**. Changes in smell after the first dose of the five most common vaccine brands **by European regions**

| Part of Europe | N | Pfizer-BioNTech | | | | | Sputnik V | | | | | AstraZeneca | | | | | Moderna | | | | | Sinopharm | | | | | Total | | | | |
|---|---|---|---|---|---|---|---|---|---|---|---|---|---|---|---|---|---|---|---|---|---|---|---|---|---|---|---|---|---|---|---|
| | | TV | SW | SW% | SI | SI% | TV | SW | SW% | SI | SI% | TV | SW | SW% | SI | SI% | TV | SW | SW% | SI | SI% | TV | SW | SW% | SI | SI% | TV | SW | SW% | SI | SI% |
| **Eastern** | **255** | 2 | 0 | 0% | 0 | 0% | 212 | 9 | 4% | 2 | 1% | 1 | 0 | 0% | 0 | 0% | 4 | 0 | 0% | 0 | 0% | 2 | 0 | 0% | 0 | 0% | **221** | **9** | **4%** | **2** | **1%** |
| **Western** | **356** | 258 | 5 | 2% | 10 | 4% | 0 | 0 | na | 0 | na | 40 | 0 | 0% | 5 | 13% | 48 | 2 | 4% | 3 | 6% | 0 | 0 | na | 0 | na | **346** | **7** | **2%** | **18** | **5%** |
| **Southern** | **219** | 147 | 1 | 1% | 4 | 3% | 0 | 0 | na | 0 | na | 28 | 2 | 7% | 1 | 4% | 23 | 1 | 4% | 5 | 22% | 0 | 0 | na | 0 | na | **198** | **4** | **2%** | **10** | **5%** |
| **Northern** | **40** | 14 | 1 | 7% | 0 | 0% | 0 | 0 | na | 0 | na | 18 | 0 | 0% | 1 | 6% | 7 | 0 | 0% | 1 | 14% | 0 | 0 | na | 0 | na | **39** | **1** | **3%** | **2** | **5%** |

N of vaccinated with the specific vaccines = 804

**Countries by regions:**

Asia – Iran. Japan. Kazakhstan. China. Taiwan. Kyrgyzstan. Afghanistan. Brunei Darussalam. Hong Kong (S.A.R.). India. Indonesia. Israel. Kuwait. Lebanon. Timor-Leste; Americas - United States of America. Canada. Mexico. Peru. Argentina. Chile. Colombia. Brazil. Ecuador; Oceania – Australia. New Zealand; Africa - South Africa. Mauritius. Algeria. Kenya. Morocco. Nigeria; Eastern Europe - Russian Federation. Bulgaria. Hungary. Ukraine. Belarus. Republic of Moldova; Western Europe – France. Germany. Switzerland. Austria. Belgium. Netherlands. Luxembourg; Northern Europe - United Kingdom of Great Britain and Northern Ireland. Sweden. Denmark. Ireland; Southern Europe – Italy. Spain.

**Supplementary Table 10. Post-vaccination changes in taste after the first dose and the first two doses of the top five common vaccine brands.**

| Brands | Vaccinated participants | | Improved taste | | Worsened taste | | Improved vs worsened[e] |
|---|---|---|---|---|---|---|---|
| | N | Frequency | N | Proportion | N | Proportion | P |
| **After the first dose of vaccine** | | | | | | | |
| **Pfizer-BioNTech**[a] | 595 | 0.49 | 13 | 2.2% | 8 | 1.3% | 0.38 |
| **Sputnik V**[b] | 245 | 0.20 | 1 | 1.4% | 5 | 2.0% | 0.22 |
| **Astra-Zeneca**[b] | 157 | 0.13 | 4 | 2.5% | 5 | 3.2% | 1 |
| **Moderna**[a] | 123 | 0.10 | 8 | 6.5% | 2 | 1.6% | 0.11 |
| **Sinopharm**[c] | 102 | 0.08 | 1 | 1.0% | 2 | 2.0% | 1 |
| **Total** | 1222 | | 27 | 2.2% | 22 | 1.8% | 0.57 |
| **Fisher's Exact test of homogeneity, p-value**[d] | | | p = 0.008 | | p = 0.561 | | |
| **After the second dose of vaccine** | | | | | | | |
| **Pfizer-BioNTech**[a] **× 2** | 528 | 0.52 | 9 | 1.7% | 5 | 0.9% | 0.42 |
| **Sputnik V**[b] **× 2** | 194 | 0.19 | 0 | 0% | 6 | 3.1% | 0.03 |
| **Astra-Zeneca**[b] **× 2** | 107 | 0.10 | 1 | 0.9% | 2 | 1.9% | 1 |
| **Moderna**[a] **× 2** | 99 | 0.10 | 6 | 6.1% | 3 | 3.0% | 0.51 |
| **Sinopharm**[c] **× 2** | 95 | 0.09 | 0 | 0% | 2 | 2.1% | 0.50 |
| **Total** | 1023 | | 16 | 1.6% | 18 | 47.1% | 0.86 |
| **Fisher's Exact test of homogeneity, p-value**[d] | | | P = 0.005 | | P = 0.16 | | |

All participants who received the first two doses of the same vaccine were included in the analysis of post-vaccination changes after the first dose of vaccine. [a]mRNA type vaccines; [b]adenovirus vector vaccines; [c]inactivated virus vaccines. [d]Fisher's test of homogeneity was used to compare the proportions of improvement or worsening between all groups. [e]Binomial tests comparing the % improvement and % worsening with two-sided p-values reported.

**Supplementary Table 11a**. Changes in taste <u>after the first dose</u> of the five most common vaccine brands **by countries**

| Country | N | Pfizer-BioNTech | | | | | Sputnik V | | | | | AstraZeneca | | | | | Moderna | | | | | Sinopharm | | | | | Total | | | | |
|---|---|---|---|---|---|---|---|---|---|---|---|---|---|---|---|---|---|---|---|---|---|---|---|---|---|---|---|---|---|---|---|
| | | TV | SW | SW% | SI | SI% | TV | SW | SW% | SI | SI% | TV | SW | SW% | SI | SI% | TV | SW | SW% | SI | SI% | TV | SW | SW% | SI | SI% | TV | SW | SW% | SI | SI% |
| **Russia** | **245** | 1 | 0 | 0% | 0 | 0% | 211 | 5 | 2% | 0 | 0% | 0 | 0 | na | 0 | na | 0 | 0 | na | 0 | na | 0 | 0 | na | 0 | na | **212** | **5** | **2%** | **0** | **0%** |
| **France** | **225** | 182 | 2 | 1% | 5 | 3% | 0 | 0 | na | 0 | na | 16 | 0 | 0% | 1 | 6% | 22 | 1 | 5% | 2 | 9% | 0 | 0 | na | 0 | na | **220** | **3** | **1%** | **8** | **4%** |
| **Iran** | **133** | 3 | 0 | 0% | 0 | 0% | 14 | 0 | 0% | 0 | 0% | 23 | 1 | 4% | 0 | 0% | 0 | 0 | na | 0 | na | 79 | 1 | 1% | 1 | 1% | **119** | **2** | **2%** | **1** | **1%** |
| **Italy** | **125** | 85 | 1 | 1% | 0 | 0% | 0 | 0 | na | 0 | na | 20 | 1 | 5% | 0 | 0% | 14 | 0 | 0% | 2 | 14% | 0 | 0 | na | 0 | na | **119** | **2** | **2%** | **2** | **2%** |
| **USA** | **112** | 58 | 1 | 2% | 2 | 3% | 0 | 0 | na | 0 | na | 1 | 0 | 0% | 0 | 0% | 34 | 1 | 3% | 2 | 6% | 0 | 0 | na | 0 | na | **93** | **2** | **2%** | **4** | **4%** |
| **Spain** | **94** | 62 | 1 | 2% | 1 | 2% | 0 | 0 | na | 0 | na | 8 | 1 | 13% | 0 | 0% | 8 | 0 | 0% | 1 | 13% | 0 | 0 | na | 0 | na | **78** | **2** | **3%** | **2** | **3%** |
| **Australia** | **92** | 61 | 0 | 0% | 0 | 0% | 0 | 0 | na | 0 | na | 25 | 0 | 0% | 0 | 0% | 1 | 0 | 0% | 0 | 0% | 1 | 0 | 0% | 0 | 0% | **88** | **0** | **0%** | **0** | **0%** |
| **Germany** | **85** | 52 | 1 | 2% | 3 | 6% | 0 | 0 | na | 0 | na | 20 | 0 | 0% | 1 | 5% | 9 | 0 | 0% | 0 | 0% | 0 | 0 | na | 0 | na | **81** | **1** | **1%** | **4** | **5%** |
| **United Kingdom** | **36** | 12 | 1 | 8% | 0 | 0% | 0 | 0 | na | 0 | na | 17 | 0 | 0% | 1 | 6% | 6 | 0 | 0% | 1 | 17% | 0 | 0 | na | 0 | na | **35** | **1** | **3%** | **2** | **6%** |
| **Canada** | **24** | 18 | 1 | 6% | 0 | 0% | 0 | 0 | na | 0 | na | 4 | 0 | 0% | 0 | 0% | 1 | 0 | 0% | 0 | 0% | 0 | 0 | na | 0 | na | **23** | **1** | **4%** | **0** | **0%** |
| **Switzerland** | **24** | 6 | 0 | 0% | 0 | 0% | 0 | 0 | na | 0 | na | 0 | 0 | na | 0 | na | 17 | 0 | 0% | 0 | 0% | 0 | 0 | na | 0 | na | **23** | **0** | **0%** | **0** | **0%** |
| **Japan** | **21** | 19 | 0 | 0% | 0 | 0% | 0 | 0 | na | 0 | na | 1 | 0 | 0% | 0 | 0% | 1 | 0 | 0% | 0 | 0% | 0 | 0 | na | 0 | na | **21** | **0** | **0%** | **0** | **0%** |
| **Kazakhstan** | **13** | 0 | 0 | na | 0 | na | 12 | 0 | 0% | 0 | 0% | 0 | 0 | na | 0 | na | 0 | 0 | na | 0 | na | 0 | 0 | na | 0 | na | **12** | **0** | **0%** | **0** | **0%** |

Countries with more than 10 participants were considered (total N = 1229. N of vaccinated with the specific vaccines = 1124); N – number of participants per country; TV – total vaccinated. N; SW. SW% - sense worsened. N. %; SI. SI% - sense improved. N. %; na – not available.

**Supplementary Table 11b.** Changes in taste <u>after the first dose</u> of the five most common vaccine brands **by world regions**

| Region | N | Pfizer-BioNTech | | | | | Sputnik V | | | | | AstraZeneca | | | | | Moderna | | | | | Sinopharm | | | | | Total | | | | |
|---|---|---|---|---|---|---|---|---|---|---|---|---|---|---|---|---|---|---|---|---|---|---|---|---|---|---|---|---|---|---|---|
| | | TV | SW | SW% | SI | SI% | TV | SW | SW% | SI | SI% | TV | SW | SW% | SI | SI% | TV | SW | SW% | SI | SI% | TV | SW | SW% | SI | SI% | TV | SW | SW% | SI | SI% |
| **Europe** | **870** | 419 | 6 | 1% | 10 | 2% | 212 | 5 | 2% | 0 | 0% | 87 | 2 | 2% | 4 | 5% | 81 | 1 | 1% | 6 | 7% | 2 | 0 | 0% | 0 | 0% | **801** | **14** | **2%** | **20** | **2%** |
| **Asia** | **193** | 28 | 0 | 0% | 0 | 0% | 27 | 0 | 0% | 0 | 0% | 30 | 2 | 7% | 0 | 0% | 2 | 0 | 0% | 0 | 0% | 83 | 2 | 2% | 1 | 1% | **170** | **4** | **2%** | **1** | **1%** |
| **Americas** | **161** | 80 | 2 | 3% | 2 | 3% | 1 | 0 | 0% | 0 | 0% | 10 | 1 | 10% | 0 | 0% | 38 | 1 | 3% | 2 | 5% | 3 | 0 | 0% | 0 | 0% | **132** | **4** | **3%** | **4** | **3%** |
| **Oceania** | **93** | 62 | 0 | 0% | 1 | 2% | 0 | 0 | na | 0 | na | 25 | 0 | 0% | 0 | 0% | 1 | 0 | 0% | 0 | 0% | 1 | 0 | 0% | 0 | 0% | **89** | **0** | **0%** | **1** | **1%** |
| **Africa** | **9** | 3 | 0 | 0% | 0 | 0% | 0 | 0 | na | 0 | na | 2 | 0 | 0% | 0 | 0% | 1 | 0 | 0% | 0 | 0% | 0 | 0 | na | 0 | na | **6** | **0** | **0%** | **0** | **0%** |

All participants who indicated their country were considered (total N = 1326. N of vaccinated with the specific vaccines = 1198)

**Supplementary Table 11c.** Changes in taste <u>after the first dose</u> of the five most common vaccine brands **by European regions**

| Part of Europe | N | Pfizer-BioNTech | | | | | Sputnik V | | | | | AstraZeneca | | | | | Moderna | | | | | Sinopharm | | | | | Total | | | | |
|---|---|---|---|---|---|---|---|---|---|---|---|---|---|---|---|---|---|---|---|---|---|---|---|---|---|---|---|---|---|---|---|
| | | TV | SW | SW% | SI | SI% | TV | SW | SW% | SI | SI% | TV | SW | SW% | SI | SI% | TV | SW | SW% | SI | SI% | TV | SW | SW% | SI | SI% | TV | SW | SW% | SI | SI% |
| **Eastern Europe** | **255** | 2 | 0 | 0% | 0 | 0% | 212 | 5 | 2% | 0 | 0% | 1 | 0 | 0% | 0 | 0% | 4 | 0 | 0% | 0 | 0% | 2 | 0 | 0% | 0 | 0% | **221** | **5** | **2%** | **0** | **0%** |
| **Western Europe** | **356** | 256 | 3 | 1% | 9 | 4% | 0 | 0 | na | 0 | na | 40 | 0 | 0% | 3 | 8% | 48 | 1 | 2% | 2 | 4% | 0 | 0 | na | 0 | na | **344** | **4** | **1%** | **14** | **4%** |
| **Southern Europe** | **219** | 147 | 2 | 1% | 1 | 1% | 0 | 0 | na | 0 | na | 28 | 2 | 7% | 0 | 0% | 22 | 0 | 0% | 3 | 14% | 0 | 0 | na | 0 | na | **197** | **4** | **2%** | **4** | **2%** |
| **Northern Europe** | **40** | 14 | 1 | 7% | 0 | 0% | 0 | 0 | na | 0 | na | 18 | 0 | 0% | 1 | 6% | 7 | 0 | 0% | 1 | 14% | 0 | 0 | na | 0 | na | **39** | **1** | **3%** | **2** | **5%** |

N of vaccinated with the specific vaccines = 801

**Countries by regions:**

Asia – Iran. Japan. Kazakhstan. China. Taiwan. Kyrgyzstan. Afghanistan. Brunei Darussalam. Hong Kong (S.A.R.). India. Indonesia. Israel. Kuwait. Lebanon. Timor-Leste; Americas - United States of America. Canada. Mexico. Peru. Argentina. Chile. Colombia. Brazil. Ecuador; Oceania – Australia. New Zealand; Africa - South Africa. Mauritius. Algeria. Kenya. Morocco. Nigeria; Eastern Europe - Russian Federation. Bulgaria. Hungary. Ukraine. Belarus. Republic of Moldova; Western Europe – France. Germany. Switzerland. Austria. Belgium. Netherlands. Luxembourg; Northern Europe - United Kingdom of Great Britain and Northern Ireland. Sweden. Denmark. Ireland; Southern Europe – Italy. Spain.

**Supplementary Table 12a.** Post-vaccination changes in pre-existing parosmia <u>after the first dose</u> by countries

| Countries | N | Pfizer-BioNTech | | | | | Moderna | | | | | Sputnik V | | | | | AstraZeneca | | | | | Total | | | | |
|---|---|---|---|---|---|---|---|---|---|---|---|---|---|---|---|---|---|---|---|---|---|---|---|---|---|---|
| | | TV | SW | SW% | SI | SI% | TV | SW | SW% | SI | SI% | TV | SW | SW% | SI | SI% | TV | SW | SW% | SI | SI% | TV | SW | SW% | SI | SI% |
| Russia | 245 | 0 | 0 | na | 0 | na | 0 | 0 | na | 0 | na | 45 | 2 | 4% | 6 | 13% | 0 | 0 | na | 0 | na | 45 | 2 | 4% | 6 | 13% |
| France | 225 | 85 | 1 | 1% | 8 | 9% | 15 | 0 | 0% | 3 | 20% | 0 | 0 | na | 0 | na | 4 | 1 | 25% | 1 | 25% | 104 | 2 | 2% | 12 | 12% |
| Iran | 133 | 0 | 0 | na | 0 | na | 0 | 0 | na | 0 | na | 1 | 0 | 0% | 0 | 0% | 0 | 0 | na | 0 | na | 1 | 0 | 0% | 0 | 0% |
| Italy | 125 | 26 | 0 | 0% | 2 | 8% | 4 | 0 | 0% | 3 | 75% | 0 | 0 | na | 0 | na | 3 | 0 | 0% | 0 | 0% | 33 | 0 | 0% | 5 | 15% |
| USA | 112 | 25 | 0 | 0% | 3 | 12% | 14 | 0 | 0% | 2 | 14% | 0 | 0 | na | 0 | na | 0 | 0 | na | 0 | na | 39 | 0 | 0% | 5 | 13% |
| Spain | 94 | 29 | 1 | 3% | 1 | 3% | 4 | 0 | 0% | 2 | 50% | 0 | 0 | na | 0 | na | 4 | 0 | 0% | 0 | 0% | 37 | 1 | 3% | 3 | 8% |
| Australia | 92 | 0 | 0 | na | 0 | na | 0 | 0 | na | 0 | na | 0 | 0 | na | 0 | na | 1 | 0 | 0% | 0 | 0% | 1 | 0 | 0% | 0 | 0% |
| Germany | 85 | 23 | 0 | 0% | 4 | 17% | 3 | 0 | 0% | 0 | 0% | 0 | 0 | na | 0 | na | 11 | 0 | 0% | 1 | 9% | 37 | 0 | 0% | 5 | 14% |
| United Kingdom | 36 | 8 | 0 | 0% | 0 | 0% | 3 | 0 | 0% | 1 | 33% | 0 | 0 | na | 0 | na | 12 | 0 | 0% | 0 | 0% | 23 | 0 | 0% | 1 | 4% |
| Canada | 24 | 11 | 1 | 9% | 1 | 9% | 0 | 0 | na | 0 | na | 0 | 0 | na | 0 | na | 3 | 0 | 0% | 1 | 33% | 14 | 1 | 7% | 2 | 14% |
| Switzerland | 24 | 1 | 0 | 0% | 0 | 0% | 6 | 0 | 0% | 1 | 17% | 0 | 0 | na | 0 | na | 0 | 0 | na | 0 | na | 7 | 0 | 0% | 1 | 14% |
| Japan | 21 | 1 | 0 | 0% | 0 | 0% | 0 | 0 | na | 0 | na | 0 | 0 | na | 0 | na | 0 | 0 | na | 0 | na | 1 | 0 | 0% | 0 | 0% |
| Kazakhstan | 13 | 0 | 0 | na | 0 | na | 0 | 0 | na | 0 | na | 6 | 2 | 33% | 2 | 33% | 0 | 0 | na | 0 | na | 6 | 2 | 33% | 2 | 33% |

Countries with more than 10 participants were considered (total N = 1229, N of vaccinated with the specific vaccines & with pre-existing parosmia = 348); N – number of participants per country; TV – total vaccinated with pre-existing parosmia, N; SW, SW% - sense worsened, N, %; SI, SI% - sense improved, N, %; na – not available.

**Supplementary Table 12b.** Post-vaccination changes in pre-existing parosmia after the first dose by world regions

| Regions | N | Pfizer-BioNTech | | | | | Moderna | | | | | Sputnik V | | | | | AstraZeneca | | | | | Total | | | | |
|---|---|---|---|---|---|---|---|---|---|---|---|---|---|---|---|---|---|---|---|---|---|---|---|---|---|---|
| | | TV | SW | SW% | SI | SI% | TV | SW | SW% | SI | SI% | TV | SW | SW% | SI | SI% | TV | SW | SW% | SI | SI% | TV | SW | SW% | SI | SI% |
| **Europe** | **870** | 182 | 3 | 2% | 15 | 8% | 38 | 0 | 0% | 11 | 29% | 46 | 2 | 4% | 6 | 13% | 37 | 1 | 3% | 2 | 5% | **303** | **6** | **2%** | **34** | **11%** |
| **Asia** | **193** | 2 | 0 | 0% | 0 | 0% | 0 | 0 | na | 0 | na | 7 | 2 | 29% | 2 | 29% | 1 | 0 | 0% | 0 | 0% | **10** | **2** | **20%** | **2** | **20%** |
| **Americas** | **161** | 36 | 1 | 3% | 4 | 11% | 15 | 0 | 0% | 2 | 13% | 0 | 0 | na | 0 | na | 6 | 1 | 17% | 1 | 17% | **57** | **2** | **4%** | **7** | **12%** |
| **Oceania** | **93** | 1 | 0 | 0% | 0 | 0% | 0 | 0 | na | 0 | na | 0 | 0 | na | 0 | na | 1 | 0 | 0% | 0 | 0% | **2** | **0** | **0%** | **0** | **0%** |
| **Africa** | **9** | 2 | 0 | 0% | 1 | 50% | 0 | 0 | na | 0 | na | 0 | 0 | na | 0 | na | 0 | 0 | na | 0 | na | **2** | **0** | **0%** | **1** | **50%** |

All participants who indicated their country were considered (total N = 1326, N of vaccinated with the specific vaccines & with pre-existing parosmia = 374)

**Supplementary Table 12c.** Post-vaccination changes in pre-existing parosmia after the first dose by European regions

| European regions | N | Pfizer-BioNTech | | | | | Moderna | | | | | Sputnik V | | | | | AstraZeneca | | | | | Total | | | | |
|---|---|---|---|---|---|---|---|---|---|---|---|---|---|---|---|---|---|---|---|---|---|---|---|---|---|---|
| | | TV | SW | SW% | SI | SI% | TV | SW | SW% | SI | SI% | TV | SW | SW% | SI | SI% | TV | SW | SW% | SI | SI% | TV | SW | SW% | SI | SI% |
| **Eastern Europe** | **255** | 1 | 0 | 0% | 0 | 0% | 2 | 0 | 0% | 0 | 0% | 46 | 2 | 4% | 6 | 13% | 0 | 0 | na | 0 | na | **49** | **2** | **4%** | **6** | **12%** |
| **Western Europe** | **356** | 117 | 2 | 2% | 12 | 10% | 24 | 0 | 0% | 4 | 17% | 0 | 0 | na | 0 | na | 18 | 1 | 6% | 2 | 11% | **159** | **3** | **2%** | **18** | **11%** |
| **Southern Europe** | **219** | 55 | 1 | 2% | 3 | 5% | 8 | 0 | 0% | 5 | 63% | 0 | 0 | na | 0 | na | 7 | 0 | 0% | 0 | 0% | **70** | **1** | **1%** | **8** | **11%** |
| **Northern Europe** | **40** | 9 | 0 | 0% | 0 | 0% | 4 | 0 | 0% | 2 | 50% | 0 | 0 | na | 0 | na | 12 | 0 | 0% | 0 | 0% | **25** | **0** | **0%** | **2** | **8%** |

N of vaccinated with the specific vaccines & with pre-existing parosmia = 303

**Countries by regions:**
Asia – Iran. Japan. Kazakhstan. China. Taiwan. Kyrgyzstan. Afghanistan. Brunei Darussalam. Hong Kong (S.A.R.). India. Indonesia. Israel. Kuwait. Lebanon. Timor-Leste; Americas - United States of America. Canada. Mexico. Peru. Argentina. Chile. Colombia. Brazil. Ecuador; Oceania – Australia. New Zealand; Africa - South Africa. Mauritius. Algeria. Kenya. Morocco. Nigeria; Eastern Europe - Russian Federation. Bulgaria. Hungary. Ukraine. Belarus. Republic of Moldova; Western Europe – France. Germany. Switzerland. Austria. Belgium. Netherlands. Luxembourg; Northern Europe - United Kingdom of Great Britain and Northern Ireland. Sweden. Denmark. Ireland; Southern Europe – Italy. Spain.

**Supplementary Table 13a.** Post-vaccination changes in pre-existing phantosmia after the first dose by countries

| Country | N | Pfizer-BioNTech | | | | | Moderna | | | | | Sputnik V | | | | | AstraZeneca | | | | | Total | | | | |
|---|---|---|---|---|---|---|---|---|---|---|---|---|---|---|---|---|---|---|---|---|---|---|---|---|---|---|
| | | TV | SW | SW% | SI | SI% | TV | SW | SW% | SI | SI% | TV | SW | SW% | SI | SI% | TV | SW | SW% | SI | SI% | TV | SW | SW% | SI | SI% |
| **Russia** | **245** | 1 | 0 | 0% | 1 | 100% | 0 | 0 | na | 0 | na | 21 | 2 | 10% | 5 | 24% | 0 | 0 | na | 0 | na | **22** | **2** | **9%** | **6** | **27%** |
| **France** | **225** | 57 | 2 | 4% | 8 | 14% | 11 | 1 | 9% | 5 | 45% | 0 | 0 | na | 0 | na | 1 | 0 | 0% | 1 | 100% | **69** | **3** | **4%** | **14** | **20%** |
| **Iran** | **133** | 0 | 0 | na | 0 | na | 0 | 0 | na | 0 | na | 0 | 0 | na | 0 | na | 0 | 0 | na | 0 | na | **0** | **0** | **na** | **0** | **na** |
| **Italy** | **125** | 15 | 0 | 0% | 5 | 33% | 3 | 1 | 33% | 1 | 33% | 0 | 0 | na | 0 | na | 4 | 0 | 0% | 2 | 50% | **22** | **1** | **5%** | **8** | **36%** |
| **USA** | **112** | 26 | 0 | 0% | 4 | 15% | 9 | 1 | 11% | 3 | 33% | 0 | 0 | na | 0 | na | 0 | 0 | na | 0 | na | **35** | **1** | **3%** | **7** | **20%** |
| **Spain** | **94** | 18 | 0 | 0% | 1 | 6% | 1 | 0 | 0% | 0 | 0% | 0 | 0 | na | 0 | na | 1 | 0 | 0% | 0 | 0% | **20** | **0** | **0%** | **1** | **5%** |
| **Australia** | **92** | 0 | 0 | na | 0 | na | 0 | 0 | na | 0 | na | 0 | 0 | na | 0 | na | 2 | 0 | 0% | 0 | 0% | **2** | **0** | **0%** | **0** | **0%** |
| **Germany** | **85** | 13 | 1 | 8% | 2 | 15% | 2 | 0 | 0% | 0 | 0% | 0 | 0 | na | 0 | na | 8 | 0 | 0% | 2 | 25% | **23** | **1** | **4%** | **4** | **17%** |
| **United Kingdom** | **36** | 5 | 0 | 0% | 0 | 0% | 2 | 0 | 0% | 1 | 50% | 0 | 0 | na | 0 | na | 7 | 0 | 0% | 1 | 14% | **14** | **0** | **0%** | **2** | **14%** |
| **Canada** | **24** | 8 | 0 | 0% | 3 | 38% | 0 | 0 | na | 0 | na | 0 | 0 | na | 0 | na | 4 | 0 | 0% | 0 | 0% | **12** | **0** | **0%** | **3** | **25%** |
| **Switzerland** | **24** | 0 | 0 | na | 0 | na | 4 | 0 | 0% | 1 | 25% | 0 | 0 | na | 0 | na | 0 | 0 | na | 0 | na | **4** | **0** | **0%** | **1** | **25%** |
| **Japan** | **21** | 0 | 0 | na | 0 | na | 0 | 0 | na | 0 | na | 0 | 0 | na | 0 | na | 0 | 0 | na | 0 | na | **0** | **0** | **na** | **0** | **na** |
| **Kazakhstan** | **13** | 0 | 0 | na | 0 | na | 0 | 0 | na | 0 | na | 2 | 0 | 0% | 1 | 50% | 0 | 0 | na | 0 | na | **2** | **0** | **0%** | **1** | **50%** |

Countries with more than 10 participants were considered (total N = 1229, N of vaccinated with the specific vaccines & with pre-existing phantosmia = 225); N – number of participants per country; TV – total vaccinated with pre-existing phantosmia, N; SW, SW% - sense worsened, N, %; SI, SI% - sense improved, N, %; na – not available.

**Supplementary Table 13b.** Post-vaccination changes in pre-existing phantosmia after the first dose by world regions

| Region | N | Pfizer-BioNTech | | | | | Moderna | | | | | Sputnik V | | | | | AstraZeneca | | | | | Total | | | | |
|---|---|---|---|---|---|---|---|---|---|---|---|---|---|---|---|---|---|---|---|---|---|---|---|---|---|---|
| | | TV | SW | SW% | SI | SI% | TV | SW | SW% | SI | SI% | TV | SW | SW% | SI | SI% | TV | SW | SW% | SI | SI% | TV | SW | SW% | SI | SI% |
| **Europe** | **870** | 114 | 3 | 3% | 18 | 16% | 23 | 2 | 9% | 8 | 35% | 21 | 2 | 10% | 5 | 24% | 23 | 0 | 0% | 6 | 26% | **181** | **7** | **4%** | **37** | **20%** |
| **Asia** | **193** | 1 | 0 | 0% | 0 | 0% | 0 | 0 | na | 0 | na | 2 | 0 | 0% | 1 | 50% | 0 | 0 | na | 0 | na | **3** | **0** | **0%** | **1** | **33%** |
| **Americas** | **161** | 34 | 0 | 0% | 7 | 21% | 9 | 1 | 11% | 3 | 33% | 0 | 0 | na | 0 | na | 5 | 0 | 0% | 0 | 0% | **48** | **1** | **2%** | **10** | **21%** |
| **Oceania** | **93** | 0 | 0 | na | 0 | na | 0 | 0 | na | 0 | na | 0 | 0 | na | 0 | na | 2 | 0 | 0% | 0 | 0% | **2** | **0** | **0%** | **0** | **0%** |
| **Africa** | **9** | 1 | 0 | 0% | 1 | 100% | 0 | 0 | na | 0 | na | 0 | 0 | na | 0 | na | 0 | 0 | na | 0 | na | **1** | **0** | **0%** | **1** | **100%** |

All participants who indicated their country were considered (total N = 1326, N of vaccinated with the specific vaccines & with pre-existing phantosmia = 235)

**Supplementary Table 13c.** Post-vaccination changes in pre-existing phantosmia after the first dose by European regions

| European regions | N | Pfizer-BioNTech | | | | | Moderna | | | | | Sputnik V | | | | | AstraZeneca | | | | | Total | | | | |
|---|---|---|---|---|---|---|---|---|---|---|---|---|---|---|---|---|---|---|---|---|---|---|---|---|---|---|
| | | TV | SW | SW% | SI | SI% | TV | SW | SW% | SI | SI% | TV | SW | SW% | SI | SI% | TV | SW | SW% | SI | SI% | TV | SW | SW% | SI | SI% |
| **Eastern Europe** | **255** | 2 | 0 | 0% | 2 | 100% | 0 | 0 | na | 0 | na | 21 | 2 | 10% | 5 | 24% | 0 | 0 | na | 0 | na | **23** | **2** | **9%** | **7** | **30%** |
| **Western Europe** | **356** | 73 | 3 | 4% | 10 | 14% | 17 | 1 | 6% | 6 | 35% | 0 | 0 | na | 0 | na | 11 | 0 | 0% | 3 | 27% | **101** | **4** | **4%** | **19** | **19%** |
| **Southern Europe** | **219** | 33 | 0 | 0% | 6 | 18% | 4 | 1 | 25% | 1 | 25% | 0 | 0 | na | 0 | na | 5 | 0 | 0% | 2 | 40% | **42** | **1** | **2%** | **9** | **21%** |
| **Northern Europe** | **40** | 6 | 0 | 0% | 0 | 0% | 2 | 0 | 0% | 1 | 50% | 0 | 0 | na | 0 | na | 7 | 0 | 0% | 1 | 14% | **15** | **0** | **0%** | **2** | **13%** |

N of vaccinated with the specific vaccines & with pre-existing phantosmia = 181

**Countries by regions:**
Asia – Iran. Japan. Kazakhstan. China. Taiwan. Kyrgyzstan. Afghanistan. Brunei Darussalam. Hong Kong (S.A.R.). India. Indonesia. Israel. Kuwait. Lebanon. Timor-Leste; Americas - United States of America. Canada. Mexico. Peru. Argentina. Chile. Colombia. Brazil. Ecuador; Oceania – Australia. New Zealand; Africa - South Africa. Mauritius. Algeria. Kenya. Morocco. Nigeria; Eastern Europe - Russian Federation. Bulgaria. Hungary. Ukraine. Belarus. Republic of Moldova; Western Europe – France. Germany. Switzerland. Austria. Belgium. Netherlands. Luxembourg; Northern Europe - United Kingdom of Great Britain and Northern Ireland. Sweden. Denmark. Ireland; Southern Europe – Italy. Spain.

**Supplementary Table 14a.** Post-vaccination changes in pre-existing distorted taste after the first dose by countries

| Country | N | Pfizer-BioNTech | | | | | Moderna | | | | | AstraZeneca | | | | | Sputnik V | | | | | Total | | | | |
|---|---|---|---|---|---|---|---|---|---|---|---|---|---|---|---|---|---|---|---|---|---|---|---|---|---|---|
| | | TV | SW | SW% | SI | SI% | TV | SW | SW% | SI | SI% | TV | SW | SW% | SI | SI% | TV | SW | SW% | SI | SI% | TV | SW | SW% | SI | SI% |
| **Russia** | **245** | 0 | 0 | na | 0 | na | 0 | 0 | na | 0 | na | 0 | 0 | na | 0 | na | 29 | 3 | 10% | 6 | 21% | **29** | **3** | **10%** | **6** | **21%** |
| **France** | **225** | 84 | 2 | 2% | 13 | 15% | 13 | 0 | 0% | 2 | 15% | 5 | 0 | 0% | 1 | 20% | 0 | 0 | na | 0 | na | **102** | **2** | **2%** | **16** | **16%** |
| **Iran** | **133** | 0 | 0 | na | 0 | na | 0 | 0 | na | 0 | na | 0 | 0 | na | 0 | na | 1 | 0 | 0% | 0 | 0% | **1** | **0** | **0%** | **0** | **0%** |
| **Italy** | **125** | 22 | 0 | 0% | 1 | 5% | 3 | 0 | 0% | 1 | 33% | 4 | 0 | 0% | 0 | 0% | 0 | 0 | na | 0 | na | **29** | **0** | **0%** | **2** | **7%** |
| **USA** | **112** | 25 | 0 | 0% | 2 | 8% | 16 | 1 | 6% | 2 | 13% | 0 | 0 | na | 0 | na | 0 | 0 | na | 0 | na | **41** | **1** | **2%** | **4** | **10%** |
| **Spain** | **94** | 30 | 1 | 3% | 2 | 7% | 2 | 1 | 50% | 1 | 50% | 2 | 0 | 0% | 0 | 0% | 0 | 0 | na | 0 | na | **34** | **2** | **6%** | **3** | **9%** |
| **Australia** | **92** | 0 | 0 | na | 0 | na | 0 | 0 | na | 0 | na | 1 | 0 | 0% | 0 | 0% | 0 | 0 | na | 0 | na | **1** | **0** | **0%** | **0** | **0%** |
| **Germany** | **85** | 18 | 1 | 6% | 2 | 11% | 3 | 0 | 0% | 0 | 0% | 9 | 0 | 0% | 1 | 11% | 0 | 0 | na | 0 | na | **30** | **1** | **3%** | **3** | **10%** |
| **United Kingdom** | **36** | 8 | 0 | 0% | 0 | 0% | 2 | 0 | 0% | 1 | 50% | 13 | 0 | 0% | 1 | 8% | 0 | 0 | na | 0 | na | **23** | **0** | **0%** | **2** | **9%** |
| **Canada** | **24** | 10 | 0 | 0% | 0 | 0% | 0 | 0 | na | 0 | na | 3 | 0 | 0% | 1 | 33% | 0 | 0 | na | 0 | na | **13** | **0** | **0%** | **1** | **8%** |
| **Switzerland** | **24** | 1 | 0 | 0% | 0 | 0% | 5 | 0 | 0% | 0 | 0% | 0 | 0 | na | 0 | na | 0 | 0 | na | 0 | na | **6** | **0** | **0%** | **0** | **0%** |
| **Japan** | **21** | 0 | 0 | na | 0 | na | 0 | 0 | na | 0 | na | 0 | 0 | na | 0 | na | 0 | 0 | na | 0 | na | **0** | **0** | **na** | **0** | **na** |
| **Kazakhstan** | **13** | 0 | 0 | na | 0 | na | 0 | 0 | na | 0 | na | 0 | 0 | na | 0 | na | 3 | 0 | 0% | 1 | 33% | **3** | **0** | **0%** | **1** | **33%** |

Countries with more than 10 participants were considered (total N = 1229, N of vaccinated with the specific vaccines & with pre-existing distorted taste = 312); N – number of participants per country; TV – total vaccinated with pre-existing distorted taste, N; SW, SW% - sense worsened, N, %; SI, SI% - sense improved, N, %; na – not available.

**Supplementary Table 14b.** Post-vaccination changes in pre-existing distorted taste after the first dose by world regions

| Region | N | Pfizer-BioNTech | | | | | Moderna | | | | | AstraZeneca | | | | | Sputnik V | | | | | Total | | | | |
|---|---|---|---|---|---|---|---|---|---|---|---|---|---|---|---|---|---|---|---|---|---|---|---|---|---|---|
| | | TV | SW | SW% | SI | SI% | TV | SW | SW% | SI | SI% | TV | SW | SW% | SI | SI% | TV | SW | SW% | SI | SI% | TV | SW | SW% | SI | SI% |
| **Europe** | **870** | 172 | 4 | 2% | 19 | 11% | 31 | 1 | 3% | 5 | 16% | 36 | 0 | 0% | 4 | 11% | 30 | 3 | 10% | 6 | 20% | **269** | **8** | **3%** | **34** | **13%** |
| **Asia** | **193** | 1 | 0 | 0% | 0 | 0% | 0 | 0 | na | 0 | na | 1 | 0 | 0% | 0 | 0% | 4 | 0 | 0% | 1 | 25% | **6** | **0** | **0%** | **1** | **17%** |
| **Americas** | **161** | 36 | 0 | 0% | 2 | 6% | 17 | 1 | 6% | 2 | 12% | 5 | 0 | 0% | 1 | 20% | 0 | 0 | na | 0 | na | **58** | **1** | **2%** | **5** | **9%** |
| **Oceania** | **93** | 1 | 0 | 0% | 1 | 100% | 0 | 0 | na | 0 | na | 1 | 0 | 0% | 0 | 0% | 0 | 0 | na | 0 | na | **2** | **0** | **0%** | **1** | **50%** |
| **Africa** | **9** | 2 | 0 | 0% | 1 | 50% | 0 | 0 | na | 0 | na | 0 | 0 | na | 0 | na | 0 | 0 | na | 0 | na | **2** | **0** | **0%** | **1** | **50%** |

All participants who indicated their country were considered (total N = 1326, N of vaccinated with the specific vaccines & with pre-existing distorted taste = 337)

**Supplementary Table 14c.** Post-vaccination changes in pre-existing distorted taste after the first dose by European regions

| European regions | N | Pfizer-BioNTech | | | | | Moderna | | | | | AstraZeneca | | | | | Sputnik V | | | | | Total | | | | |
|---|---|---|---|---|---|---|---|---|---|---|---|---|---|---|---|---|---|---|---|---|---|---|---|---|---|---|
| | | TV | SW | SW% | SI | SI% | TV | SW | SW% | SI | SI% | TV | SW | SW% | SI | SI% | TV | SW | SW% | SI | SI% | TV | SW | SW% | SI | SI% |
| **Eastern Europe** | **255** | 0 | 0 | na | 0 | na | 2 | 0 | 0% | 0 | 0% | 1 | 0 | 0% | 0 | 0% | 30 | 3 | 10% | 6 | 20% | **33** | **3** | **9%** | **6** | **18%** |
| **Western Europe** | **356** | 112 | 3 | 3% | 16 | 14% | 21 | 0 | 0% | 2 | 10% | 16 | 0 | 0% | 3 | 19% | 0 | 0 | na | 0 | na | **149** | **3** | **2%** | **21** | **14%** |
| **Southern Europe** | **219** | 52 | 1 | 2% | 3 | 6% | 5 | 1 | 20% | 2 | 40% | 6 | 0 | 0% | 0 | 0% | 0 | 0 | na | 0 | na | **63** | **2** | **3%** | **5** | **8%** |
| **Northern Europe** | **40** | 8 | 0 | 0% | 0 | 0% | 3 | 0 | 0% | 1 | 33% | 13 | 0 | 0% | 1 | 8% | 0 | 0 | na | 0 | na | **24** | **0** | **0%** | **2** | **8%** |

N of vaccinated with the specific vaccines & with pre-existing distorted taste = 269

**Countries by regions:**

Asia – Iran. Japan. Kazakhstan. China. Taiwan. Kyrgyzstan. Afghanistan. Brunei Darussalam. Hong Kong (S.A.R.). India. Indonesia. Israel. Kuwait. Lebanon. Timor-Leste; Americas - United States of America. Canada. Mexico. Peru. Argentina. Chile. Colombia. Brazil. Ecuador; Oceania – Australia. New Zealand; Africa - South Africa. Mauritius. Algeria. Kenya. Morocco. Nigeria; Eastern Europe - Russian Federation. Bulgaria. Hungary. Ukraine. Belarus. Republic of Moldova; Western Europe – France. Germany. Switzerland. Austria. Belgium. Netherlands. Luxembourg; Northern Europe - United Kingdom of Great Britain and Northern Ireland. Sweden. Denmark. Ireland; Southern Europe – Italy. Spain.

**Supplementary Table 15. Counts of self-reported side effects from COVID-19 vaccination by doses.**

| Side effect | Dose 1 | Dose 2 | Booster 1 | Booster 2 |
|---|---|---|---|---|
| fatigue | 454 (36%) | 394 (31%) | 257 (29%) | 71 (26%) |
| headache | 417 (31%) | 332 (26%) | 206 (23%) | 45 (17%) |
| fever | 428 (32%) | 297 (23%) | 199 (22%) | 40 (15%) |
| muscle pain | 387 (29%) | 305 (24%) | 206 (23%) | 50 (19%) |
| chills | 330 (24%) | 242 (19%) | 165 (18%) | 43 (16%) |
| joint pain | 295 (22%) | 224 (18%) | 149 (17%) | 36 (13%) |
| brain fog | 78 (6%) | 57 (4%) | 43 (5%) | 13 (5%) |
| other | 72 (5%) | 58 (5%) | 44 (5%) | 13 (4%) |
| sleep disturbance | 71 (5%) | 56 (4%) | 45 (5%) | 17 (6%) |
| lightheadedness | 74 (5%) | 61 (5%) | 45 (5%) | 9 (3%) |
| dizziness | 78 (6%) | 61 (5%) | 33 (4%) | 9 (3%) |
| menstrual change* | 67 (10%) | 54 (9%) | 36 (8%) | 5 (4%) |
| shortness of breath | 43 (3%) | 61 (5%) | 24 (3%) | 7 (3%) |
| nausea | 48 (3%) | 39 (3%) | 24 (3%) | 8 (3%) |
| nasal congestion | 45 (3%) | 38 (3%) | 26 (3%) | 4 (1%) |
| sore throat | 39 (3%) | 31 (2%) | 23 (3%) | 4 (1%) |
| cough | 24 (2%) | 24 (2%) | 19 (2%) | 6 (2%) |
| stomachache | 24 (2%) | 26 (2%) | 12 (1%) | 6 (2%) |
| diarrhea | 34 (3%) | 17 (1%) | 12 (1%) | 3 (1%) |
| heart problems | 22 (2%) | 22 (2%) | 14 (2%) | 4 (1%) |
| skin rash | 21 (2%) | 17 (1%) | 5 (0.6%) | 1 (0.3%) |
| dry nose | 19 (1%) | 15 (1%) | 8 (0.9%) | 1 (0.3%) |
| vomit | 13 (1%) | 9 (0.7%) | 5 (0.6%) | 4 (1%) |
| blood clots | 2 (0.1%) | 0 | 1 (0.1%) | 0 |

*Frequencies of the menstrual change were calculated in cis women.

**Supplementary Table 16. Significance level (p-value) for the k-proportions test with Marascuilo procedure testing the difference in incidence rate for each side effect across the first three doses of vaccine of the same type.**

| Vaccine type | Fever | Headache | Chills | Joint pain | Muscle pain | Fatigue |
|---|---|---|---|---|---|---|
| mRNA | 0.843 | 0.923 | 0.687 | 0.941 | 0.864 | 0.429 |
| Adenovirus | 0.000 | 0.169 | 0.019 | 0.431 | 0.559 | 0.394 |
| Inactivated virus | 0.212 | 0.031 | 0.210 | 0.907 | 0.676 | 0.423 |
| Mixed | <0.0001 | 0.028 | 0.004 | 0.004 | 0.017 | 0.100 |

**Supplementary Table 17. Incidence rates of side effects for those significantly differing between the first three doses of vaccines (p < 0.05 from Supplementary Table 13).**

| Vaccine type | Side effect | Dose 1 | Dose 2 | Booster 1 |
|---|---|---|---|---|
| Adenovirus | Fever | 0.538[b] | 0.363[ab] | 0.238[a] |
| Adenovirus | Chills | 0.375[b] | 0.200[a] | 0.213[ab] |
| Inactivated virus | Headache | 0.235[b] | 0.059[a] | 0.118[ab] |
| Mixed | Fever | 0.413[b] | 0.209[a] | 0.221[a] |
| Mixed | Headache | 0.337[b] | 0.267[ab] | 0.209[a] |
| Mixed | Chills | 0.285[b] | 0.151[a] | 0.169[a] |
| Mixed | Joint pain | 0.279[b] | 0.145[a] | 0.169[a] |
| Mixed | Muscle pain | 0.343[b] | 0.238[ab] | 0.215[a] |

[a,b] Whithin row, incidence rates (proportions) with a common letter are not significantly different (p > 0.05).

**Supplementary Table 18. Frequency of individual side effects by post-vaccination changes in smell and taste.**

| CHANGES IN GENERAL SMELL | | | | | | | | | | | | | | | | |
|---|---|---|---|---|---|---|---|---|---|---|---|---|---|---|---|---|
| | After Dose 1 | | | | After Dose 2 | | | | After Booster 1 | | | | After Booster 2 | | | |
| Side effects | worsened | improved | no change | total | worsened | improved | no change | total | worsened | improved | no change | total | worsened | improved | no change | total |
| **Fatigue** | 17 (4%) | 14 (3%) | 392 (93%) | 423 | 10 (3%) | 11 (3%) | 345 (94%) | 366 | 6 (3%) | 13 (5%) | 218 (92%) | 237 | 3 (5%) | 2 (3%) | 57 (92%) | 62 |
| **Fever** | 21 (5%) | 16 (4%) | 365 (91%) | 402 | 11 (4%) | 6 (2%) | 255 (94%) | 272 | 4 (2%) | 7 (4%) | 173 (94%) | 184 | 2 (6%) | 1 (3%) | 30 (91%) | 33 |
| **Headache** | 16 (4%) | 14 (4%) | 355 (92%) | 385 | 8 (3%) | 11 (4%) | 291 (94%) | 310 | 3 (2%) | 7 (4%) | 180 (95%) | 190 | 3 (8%) | 0 (0%) | 36 (92%) | 39 |
| **Muscle pain** | 18 (5%) | 11 (3%) | 331 (92%) | 360 | 10 (4%) | 7 (2%) | 268 (94%) | 285 | 6 (3%) | 6 (3%) | 184 (94%) | 196 | 3 (6%) | 3 (6%) | 41 (87%) | 47 |
| **Chills** | 16 (5%) | 16 (5%) | 282 (90%) | 314 | 10 (4%) | 7 (3%) | 211 (93%) | 228 | 3 (2%) | 6 (4%) | 144 (94%) | 153 | 1 (3%) | 1 (3%) | 35 (95%) | 37 |
| **Joint ache** | 14 (5%) | 10 (4%) | 253 (91%) | 277 | 7 (3%) | 8 (4%) | 189 (93%) | 204 | 6 (4%) | 5 (4%) | 127 (92%) | 138 | 1 (3%) | 2 (6%) | 29 (91%) | 32 |
| **Brain fog** | 6 (8%) | 6 (8%) | 61 (84%) | 73 | 8 (16%) | 2 (4%) | 40 (80%) | 50 | 2 (5%) | 4 (11%) | 32 (84%) | 38 | 1 (9%) | 0 (0%) | 10 (91%) | 11 |
| **Other** | 7 (10%) | 5 (7%) | 58 (83%) | 70 | 7 (13%) | 1 (2%) | 47 (85%) | 55 | 3 (7%) | 1 (2%) | 37 (90%) | 41 | 0 (0%) | 0 (0%) | 10 (100%) | 10 |
| **Lightheadedness** | 4 (6%) | 4 (6%) | 60 (88%) | 68 | 4 (7%) | 2 (4%) | 50 (89%) | 56 | 1 (2%) | 2 (5%) | 38 (93%) | 41 | 0 (0%) | 0 (0%) | 8 (100%) | 8 |
| **Dizziness** | 8 (12%) | 3 (4%) | 57 (84%) | 68 | 6 (11%) | 1 (2%) | 50 (88%) | 57 | 1 (4%) | 2 (8%) | 22 (88%) | 25 | 1 (17%) | 0 (0%) | 5 (83%) | 6 |
| **Sleep disturbances** | 6 (9%) | 6 (9%) | 54 (82%) | 66 | 4 (8%) | 3 (6%) | 45 (87%) | 52 | 5 (13%) | 2 (5%) | 33 (83%) | 40 | 2 (15%) | 0 (0%) | 11 (85%) | 13 |
| **Menstrual change** | 3 (5%) | 1 (2%) | 61 (94%) | 65 | 2 (4%) | 2 (4%) | 48 (92%) | 52 | 1 (3%) | 1 (3%) | 31 (94%) | 33 | 1 (20%) | 0 (0%) | 4 (80%) | 5 |
| **Nausea** | 6 (13%) | 0 (0%) | 40 (87%) | 46 | 5 (13%) | 1 (3%) | 33 (85%) | 39 | 2 (9%) | 2 (9%) | 19 (83%) | 23 | 2 (33%) | 0 (0%) | 4 (67%) | 6 |
| **Nasal congestion** | 5 (12%) | 2 (5%) | 34 (83%) | 41 | 5 (14%) | 0 (0%) | 32 (86%) | 37 | 5 (20%) | 0 (0%) | 20 (80%) | 25 | 0 (0%) | 0 (0%) | 1 (100%) | 1 |
| **Shortness of breath** | 5 (13%) | 2 (5%) | 31 (82%) | 38 | 4 (13%) | 1 (3%) | 26 (84%) | 31 | 2 (10%) | 1 (5%) | 17 (85%) | 20 | 1 (20%) | 0 (0%) | 4 (80%) | 5 |
| **Sore throat** | 6 (17%) | 1 (3%) | 29 (81%) | 36 | 5 (18%) | 1 (4%) | 22 (79%) | 28 | 3 (14%) | 1 (5%) | 17 (81%) | 21 | 1 (33%) | 0 (0%) | 2 (67%) | 3 |
| **Diarrhea** | 4 (13%) | 2 (6%) | 26 (81%) | 32 | 2 (13%) | 0 (0%) | 14 (88%) | 16 | 1 (9%) | 0 (0%) | 10 (91%) | 11 | 1 (50%) | 0 (0%) | 1 (50%) | 2 |
| **Stomach ache** | 2 (9%) | 2 (9%) | 19 (83%) | 23 | 2 (8%) | 1 (4%) | 21 (88%) | 24 | 1 (10%) | 0 (0%) | 9 (90%) | 10 | 1 (25%) | 0 (0%) | 3 (75%) | 4 |
| **Cough** | 4 (17%) | 1 (4%) | 18 (78%) | 23 | 2 (9%) | 0 (0%) | 21 (91%) | 23 | 4 (21%) | 1 (5%) | 14 (74%) | 19 | 1 (25%) | 0 (0%) | 3 (75%) | 4 |
| **Heart problems** | 6 (29%) | 2 (10%) | 13 (62%) | 21 | 5 (24%) | 1 (5%) | 15 (71%) | 21 | 1 (8%) | 0 (0%) | 11 (92%) | 12 | 1 (33%) | 0 (0%) | 2 (67%) | 3 |
| **Dry/burning nose** | 6 (32%) | 2 (11%) | 11 (58%) | 19 | 7 (50%) | 1 (7%) | 6 (43%) | 14 | 2 (25%) | 1 (13%) | 5 (63%) | 8 | | | | 0 |
| **Skin rash** | 4 (25%) | 1 (6%) | 11 (69%) | 16 | 2 (17%) | 0 (0%) | 10 (83%) | 12 | 1 (25%) | 0 (0%) | 3 (75%) | 4 | 1 (100%) | 0 (0%) | 0 (0%) | 1 |
| **Vomit** | 2 (18%) | 0 (0%) | 9 (82%) | 11 | 2 (22%) | 0 (0%) | 7 (78%) | 9 | 1 (20%) | 0 (0%) | 4 (80%) | 5 | 1 (50%) | 0 (0%) | 1 (50%) | 2 |
| **Blood clots** | 0 (0%) | 0 (0%) | 1 (100%) | 1 | | | | 0 | 0 (0%) | 0 (0%) | 1 (100%) | 1 | | | | 0 |

| CHANGES IN GENERAL TASTE | | | | | | | | | | | | | | | | |
|---|---|---|---|---|---|---|---|---|---|---|---|---|---|---|---|---|
| | After Dose 1 | | | | After Dose 2 | | | | After Booster 1 | | | | After Booster 2 | | | |
| Side effects | worsened | improved | no change | total | worsened | improved | no change | total | worsened | improved | no change | total | worsened | improved | no change | total |
| Fatigue | 11 (3%) | 11 (3%) | 398 (95%) | 420 | 8 (2%) | 5 (1%) | 351 (96%) | 364 | 3 (1%) | 8 (3%) | 227 (95%) | 238 | 2 (3%) | 2 (3%) | 60 (94%) | 64 |
| Fever | 13 (3%) | 12 (3%) | 371 (94%) | 396 | 9 (3%) | 7 (3%) | 249 (94%) | 265 | 1 (1%) | 6 (3%) | 173 (96%) | 180 | 1 (3%) | 1 (3%) | 32 (94%) | 34 |
| Headache | 10 (3%) | 13 (3%) | 357 (94%) | 380 | 5 (2%) | 7 (2%) | 292 (96%) | 304 | 1 (1%) | 4 (2%) | 181 (97%) | 186 | 2 (5%) | 0 (0%) | 37 (95%) | 39 |
| Muscle pain | 10 (3%) | 11 (3%) | 336 (94%) | 357 | 7 (2%) | 5 (2%) | 269 (96%) | 281 | 3 (2%) | 5 (3%) | 185 (96%) | 193 | 2 (4%) | 3 (6%) | 42 (89%) | 47 |
| Chills | 9 (3%) | 12 (4%) | 282 (93%) | 303 | 6 (3%) | 4 (2%) | 208 (95%) | 218 | 1 (1%) | 4 (3%) | 144 (97%) | 149 | 0 (0%) | 1 (3%) | 36 (97%) | 37 |
| Joint ache | 12 (4%) | 9 (3%) | 250 (92%) | 271 | 7 (3%) | 7 (3%) | 187 (93%) | 201 | 4 (3%) | 5 (4%) | 128 (93%) | 137 | 1 (3%) | 2 (6%) | 29 (91%) | 32 |
| Brain fog | 4 (6%) | 5 (7%) | 63 (88%) | 72 | 3 (6%) | 2 (4%) | 46 (90%) | 51 | 1 (3%) | 2 (5%) | 37 (93%) | 40 | 0 (0%) | 0 (0%) | 11 (100%) | 11 |
| Dizziness | 6 (9%) | 3 (4%) | 58 (87%) | 67 | 3 (5%) | 1 (2%) | 51 (93%) | 55 | 1 (4%) | 2 (7%) | 25 (89%) | 28 | 1 (14%) | 0 (0%) | 6 (86%) | 7 |
| Other | 5 (7%) | 3 (4%) | 59 (88%) | 67 | 8 (15%) | 0 (0%) | 46 (85%) | 54 | 1 (2%) | 1 (2%) | 39 (95%) | 41 | 0 (0%) | 0 (0%) | 11 (100%) | 11 |
| Lightheadedness | 2 (3%) | 4 (6%) | 60 (91%) | 66 | 1 (2%) | 1 (2%) | 52 (96%) | 54 | 1 (2%) | 1 (2%) | 40 (95%) | 42 | 0 (0%) | 0 (0%) | 8 (100%) | 8 |
| Sleep disturbances | 3 (5%) | 5 (8%) | 54 (87%) | 62 | 2 (4%) | 3 (6%) | 46 (90%) | 51 | 2 (5%) | 1 (3%) | 37 (93%) | 40 | 1 (7%) | 0 (0%) | 13 (93%) | 14 |
| Menstrual change | 1 (2%) | 0 (0%) | 58 (98%) | 59 | 3 (7%) | 0 (0%) | 43 (93%) | 46 | 0 (0%) | 0 (0%) | 31 (100%) | 31 | 0 (0%) | 0 (0%) | 4 (100%) | 4 |
| Nausea | 3 (7%) | 0 (0%) | 41 (93%) | 44 | 4 (11%) | 1 (3%) | 33 (87%) | 38 | 1 (4%) | 1 (4%) | 21 (91%) | 23 | 2 (29%) | 0 (0%) | 5 (71%) | 7 |
| Nasal congestion | 3 (8%) | 2 (5%) | 34 (87%) | 39 | 2 (5%) | 0 (0%) | 35 (95%) | 37 | 2 (8%) | 0 (0%) | 22 (92%) | 24 | 0 (0%) | 0 (0%) | 2 (100%) | 2 |
| Shortness of breath | 2 (6%) | 2 (6%) | 32 (89%) | 36 | 2 (7%) | 1 (3%) | 27 (90%) | 30 | 1 (5%) | 0 (0%) | 20 (95%) | 21 | 1 (20%) | 0 (0%) | 4 (80%) | 5 |
| Sore throat | 3 (9%) | 1 (3%) | 30 (88%) | 34 | 2 (7%) | 0 (0%) | 26 (93%) | 28 | 1 (5%) | 0 (0%) | 19 (95%) | 20 | 1 (33%) | 0 (0%) | 2 (67%) | 3 |
| Diarrhea | 2 (6%) | 2 (6%) | 28 (88%) | 32 | 2 (13%) | 0 (0%) | 13 (87%) | 15 | 1 (10%) | 0 (0%) | 9 (90%) | 10 | 1 (50%) | 0 (0%) | 1 (50%) | 2 |
| Stomach ache | 1 (4%) | 2 (9%) | 20 (87%) | 23 | 1 (4%) | 1 (4%) | 21 (91%) | 23 | 0 (0%) | 0 (0%) | 11 (100%) | 11 | 1 (20%) | 0 (0%) | 4 (80%) | 5 |
| Cough | 1 (5%) | 1 (5%) | 19 (90%) | 21 | 1 (5%) | 0 (0%) | 21 (95%) | 22 | 2 (11%) | 0 (0%) | 16 (89%) | 18 | 1 (25%) | 0 (0%) | 3 (75%) | 4 |
| Heart problems | 2 (10%) | 2 (10%) | 16 (80%) | 20 | 3 (15%) | 1 (5%) | 16 (80%) | 20 | 1 (8%) | 0 (0%) | 12 (92%) | 13 | 0 (0%) | 0 (0%) | 3 (100%) | 3 |
| Dry/burning nose | 4 (24%) | 3 (18%) | 10 (59%) | 17 | 4 (29%) | 1 (7%) | 9 (64%) | 14 | 2 (25%) | 0 (0%) | 6 (75%) | 8 | | | | 0 |
| Skin rash | 1 (7%) | 1 (7%) | 13 (87%) | 15 | 0 (0%) | 0 (0%) | 11 (100%) | 11 | 0 (0%) | 0 (0%) | 4 (100%) | 4 | 1 (100%) | 0 (0%) | 0 (0%) | 1 |
| Vomit | 2 (22%) | 0 (0%) | 7 (78%) | 9 | 2 (25%) | 0 (0%) | 6 (75%) | 8 | 0 (0%) | 0 (0%) | 5 (100%) | 5 | 1 (33%) | 0 (0%) | 2 (67%) | 3 |
| Blood clots | 0 (0%) | 0 (0%) | 1 (100%) | 1 | | | | 0 | 0 (0%) | 0 (0%) | 1 (100%) | 1 | | | | 0 |

### Supplementary Figures

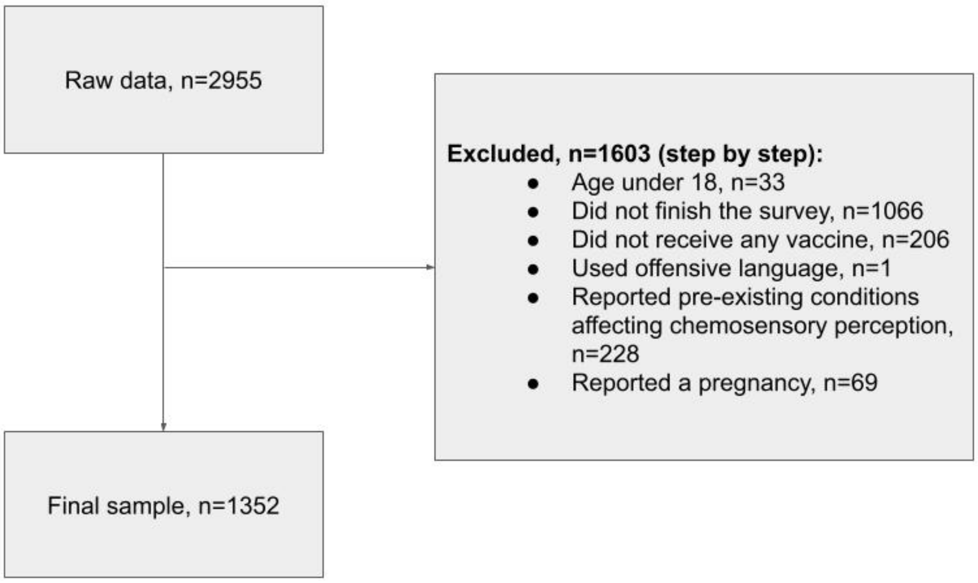


**Supplementary Figure 1. Flow diagram describing the exclusion criteria and the number of participants excluded and included in the current study**

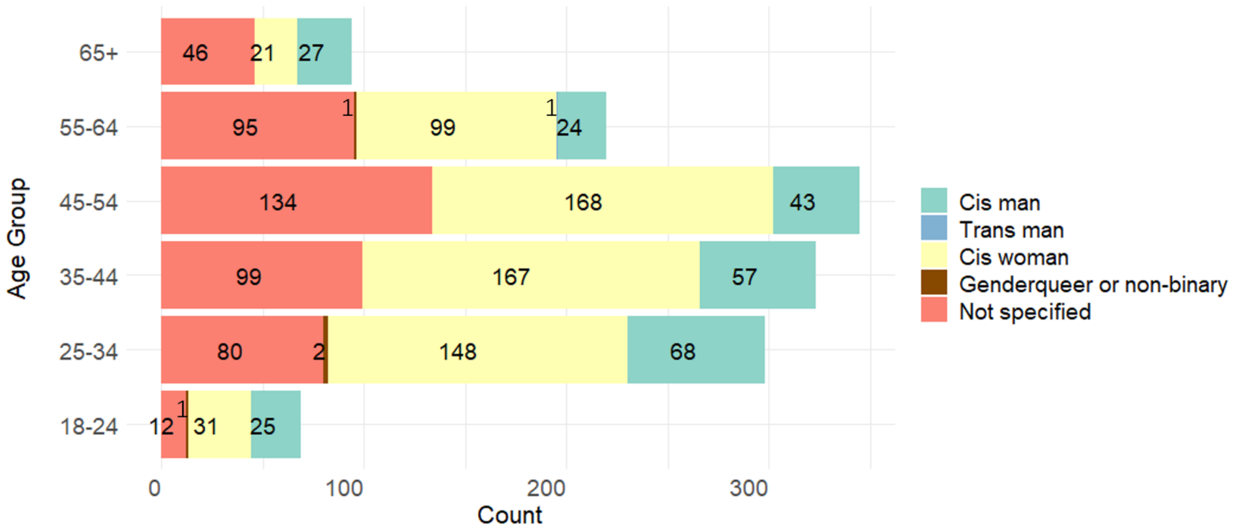


**Supplementary Figure 2. Age and gender identity of the participants.** 1349 of 1352 participants reported their gender or sex orientation.

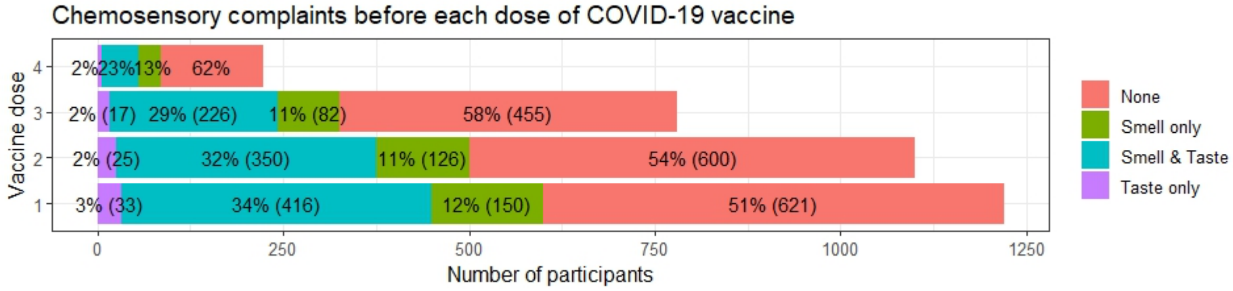


**Supplementary Figure 3. Chemosensory impairments due to COVID-19 as reported before each dose of vaccine.** Chemosensory complaints include uncovered impaired ability to smell and/or taste, parosmia, phantosmia, and distorted taste due to COVID-19. The number of participants per condition for the first three doses is shown in parentheses. To ensure unambiguous classification of chemosensory dysfunction, only participants with definitive answers for both modalities were included. The numbers of removed responses are 132, 141, 85, and 21 for doses 1-4 respectively.

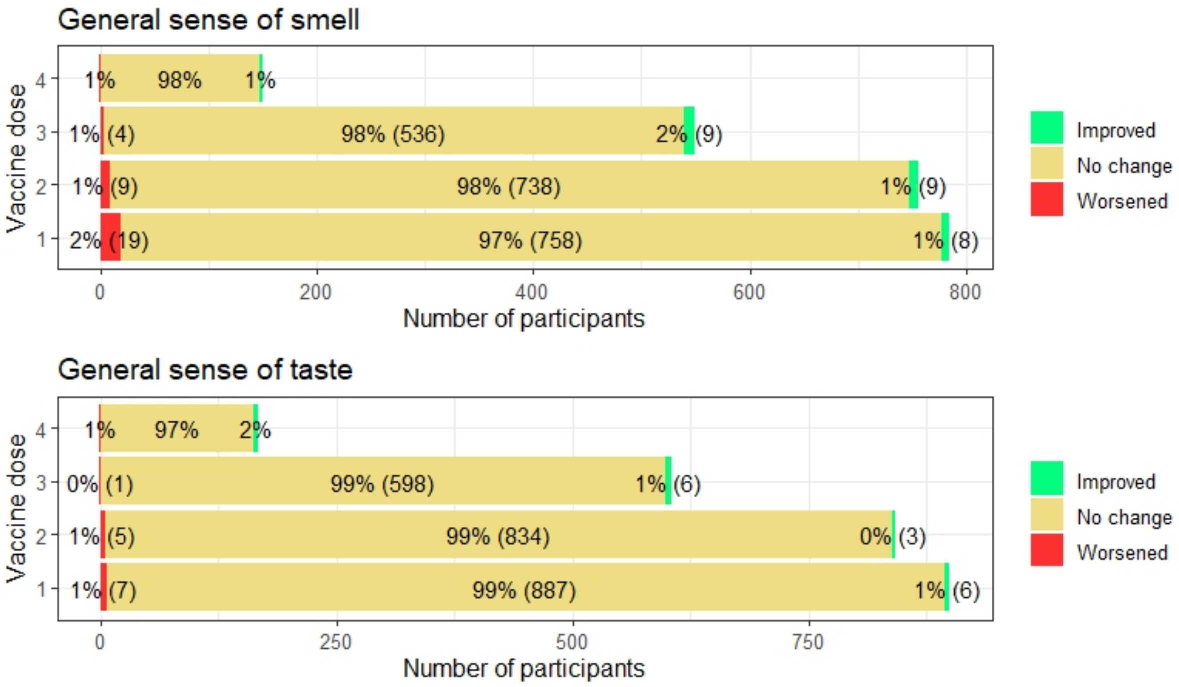


**Supplementary Figure 4. Changes in the senses of smell and taste after each dose of vaccine in participants without pre-existing sensory impairment due to COVID-19.** The number of participants per condition for the first three doses is shown in parentheses.

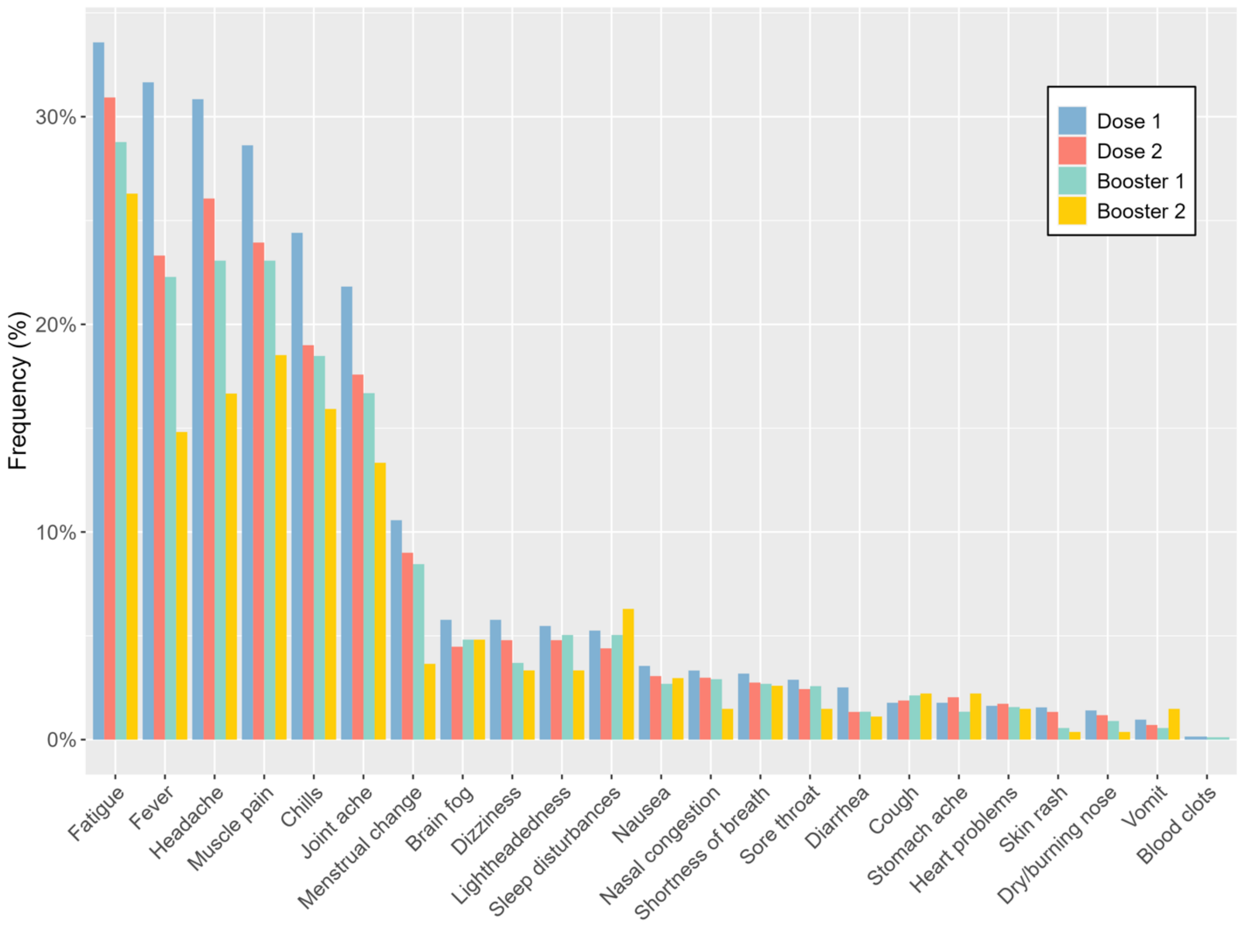


**Supplementary Figure 5. Frequency of self-reported side effects by vaccination doses.** Frequencies of the menstrual change were calculated in cis women.

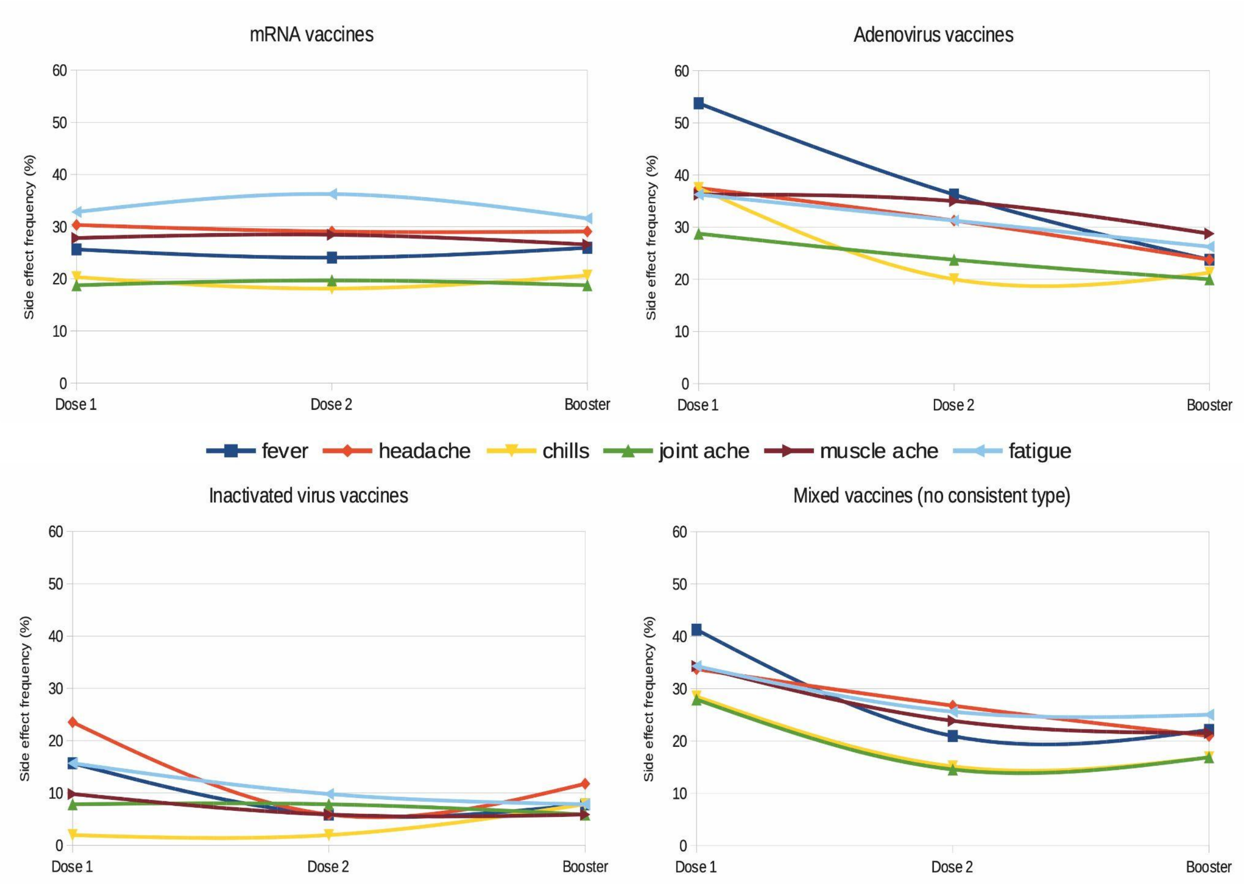


**Supplementary Figure 6. Frequencies of adverse side effects after COVID-19 vaccination for four types of vaccines.** There are 320 individuals in the mRNA vaccine group, 80 in the adenovirus vaccine group, 51 in the inactivated virus vaccine group, and 172 in the mixed vaccine group.

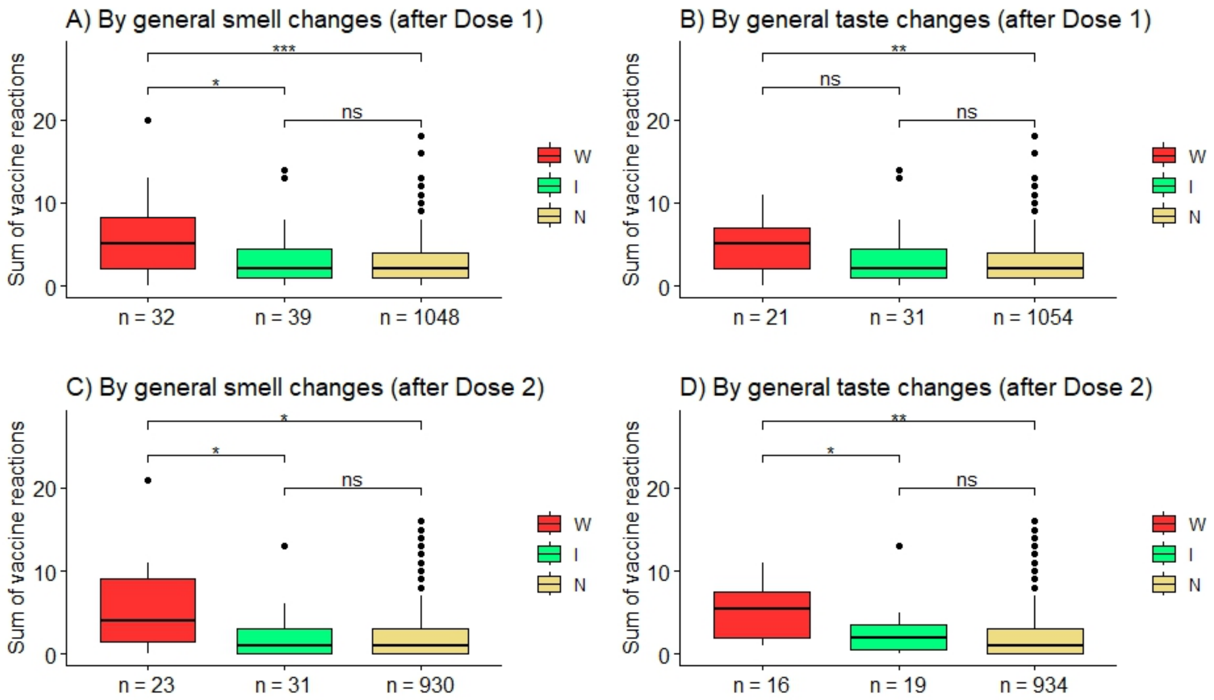


**Supplementary Figure 7. Numbers of post-vaccination side effects in groups defined by post-vaccination changes in smell and taste.** W, worsened; I, improved; N, no change. Pairwise t-test p-values are shown above brackets: ***$p < 0.001$, **$p < 0.01$, *$p < 0.05$, ns, not significant. One-way ANOVA results: A. $F(1,1117) = 47.59$, $p < 0.001$, eta squared ($\eta^2$) = 0.041; B. $F(1,1104) = 25.82$, $p < 0.001$, eta squared ($\eta^2$) = 0.023; C. $F(1, 982) = 28.11$, $p < 0.001$, eta squared ($\eta^2$) = 0.028; D. $F(1, 967) = 20.13$, $p < 0.001$, eta squared ($\eta^2$) = 0.02

GCCR

Global Consortium for Chemosensory Research

**Welcome and consent**

**Welcome and Consent**

Since the deployment of COVID-19 vaccines in autumn 2020, 11.4 billion COVID-19 vaccine doses having been administered worldwide as of today (12 April 2022). Various case reports indicate that patients with loss of smell and taste perceived a change in their ability to smell and taste after vaccination.

We would like to investigate post-vaccination changes in the perception of smell and taste and other symptoms of long-COVID. The results will help assess the potential impact of COVID-19 vaccines and provide new clues to help us understand recovery from long-COVID.

You will be asked about your experience following the vaccination in this one-time survey. It will take about 10 minutes or less to complete. Your participation is voluntary. There is a potential risk of losing confidentiality and possibly feeling uncomfortable answering various survey questions. The data will be de-identified, saved and protected in our system. You can choose to not answer any question that make you feel uncomfortable. By completing this survey, you are consenting to participate in this study. Before starting, please have your vaccine details ready.

Please let us know if you have any questions regarding the study.

General contact:
Dr Daniel Hwang
Institute for Molecular Bioscience
The University of Queensland
+61-7-3346-2630

Dr Sachiko Koyama
Indiana University
+1-812-345-6155


I am at least 18 years old, and I voluntarily participate in the present study.

- Yes
- No

**Vaccination**

**About your COVID-19 vaccination status**

What is your vaccination status?

- I did not receive any COVID-19 vaccine
- I received 1 dose
- I received 2 doses
- I received 2 doses and 1 booster
- I received more than 1 booster

When did you receive your vaccination and what is the type of the vaccine? If you received one-dose vaccines (such as Johnson & Johnson) and did not receive dose 2 before your booster shot, please skip dose 2. For doses you did not receive, please leave them blank.

| | Month | Year | Type of vaccine |
|---|---|---|---|
| Dose 1 | | | |
| Dose 2 | | | |
| Booster | | | |
| Booster 2 | | | |

If you selected other, feel free to use this space to share more information.

**About your reaction to the COVID-19 vaccine**

Did you experience any reaction? Please just provide answers to the doses you received.

| | Please answer by the dose of vaccine | | | |
|---|---|---|---|---|
| | Dose 1 | Dose 2 | Booster | Booster 2 |
| Fever | ☐ | ☐ | ☐ | ☐ |
| Headache | ☐ | ☐ | ☐ | ☐ |
| Chills | ☐ | ☐ | ☐ | ☐ |
| Diarrhea | ☐ | ☐ | ☐ | ☐ |
| Vomit | ☐ | ☐ | ☐ | ☐ |
| Stomach ache | ☐ | ☐ | ☐ | ☐ |
| Joint ache | ☐ | ☐ | ☐ | ☐ |
| Cough | ☐ | ☐ | ☐ | ☐ |
| Sore throat | ☐ | ☐ | ☐ | ☐ |
| Shortness of breath | ☐ | ☐ | ☐ | ☐ |
| Muscle soreness/pain other than at the site of injection | ☐ | ☐ | ☐ | ☐ |
| Lightheadedness | ☐ | ☐ | ☐ | ☐ |
| Dizziness | ☐ | ☐ | ☐ | ☐ |
| Nausea | ☐ | ☐ | ☐ | ☐ |
| Fatigue | ☐ | ☐ | ☐ | ☐ |
| Nasal congestion | ☐ | ☐ | ☐ | ☐ |
| Dry/burning nose | ☐ | ☐ | ☐ | ☐ |
| Heart problems | ☐ | ☐ | ☐ | ☐ |
| Sleep disturbances | ☐ | ☐ | ☐ | ☐ |
| Blood clots | ☐ | ☐ | ☐ | ☐ |
| Skin rash | ☐ | ☐ | ☐ | ☐ |
| Menstral change | ☐ | ☐ | ☐ | ☐ |
| Brain fog | ☐ | ☐ | ☐ | ☐ |
| Other (Please use the space below to specify) | ☐ | ☐ | ☐ | ☐ |
| None of above | ☐ | ☐ | ☐ | ☐ |
| I do not remember | ☐ | ☐ | ☐ | ☐ |

If you selected other, feel free to use this space to share more information.

If you had any reaction to the vaccine, how long did the reaction last? Please answer by the dose of vaccine you received.

| | Number of days |
|---|---|
| Dose 1 | |
| Dose 2 | |
| Booster | |
| Booster 2 | |

Did you get medicine or medical treatment after vaccination? Please answer by the dose of vaccine you received.

| | Select your answer from the drop down list |
|---|---|
| Dose 1 | |
| Dose 2 | |
| Booster | |
| Booster 2 | |

If you selected other, feel free to use this space to share more information.

**This section is about your sense of SMELL**

How was your sense of smell before vaccination? Please answer by the dose of vaccine you received.

| | Select your answer from the drop down list |
|---|---|
| Dose 1 | |
| Dose 2 | |
| Booster | |
| Booster 2 | |

If you selected other, feel free to use this space to share more information.

Did the vaccination affect your sense of smell? Please answer by the dose of vaccine you received.

| | Yes, sense worsened | Yes, sense improved | No | Not applicable |
|---|---|---|---|---|
| Dose 1 | ○ | ○ | ○ | ○ |
| Dose 2 | ○ | ○ | ○ | ○ |
| Booster | ○ | ○ | ○ | ○ |
| Booster 2 | ○ | ○ | ○ | ○ |

If vaccination affected your sense of smell, when did the change start? Please answer by the dose of vaccine you received.

| | Immediately after vaccination | Within 1 day of vaccination | After 1 day of vaccination | The vaccination did not affect my sense of smell |
|---|---|---|---|---|
| Dose 1 | ○ | ○ | ○ | ○ |
| Dose 2 | ○ | ○ | ○ | ○ |
| Booster | ○ | ○ | ○ | ○ |
| Booster 2 | ○ | ○ | ○ | ○ |

If vaccination affected your sense of smell, how long did the change last? Please answer by the dose of vaccine you received.

| | <1 day | 1 day to 1 week | 1 week to 1 month | >1 month | No effect |
|---|---|---|---|---|---|
| Dose 1 | ○ | ○ | ○ | ○ | ○ |
| Dose 2 | ○ | ○ | ○ | ○ | ○ |
| Booster | ○ | ○ | ○ | ○ | ○ |
| Booster 2 | ○ | ○ | ○ | ○ | ○ |

**Parosmia**, or distorted smell, occurs when things smell differently than they did before. For example, a person would have parosmia if their morning coffee now smells like gasoline.

Did you experience parosmia after your vaccination? Please answer by the dose of vaccine you received.

| | Select your answer from the drop down list |
|---|---|
| Dose 1 | [ ] |

| | Select your answer from the drop down list |
|---|---|
| Dose 2 | |
| Booster | |
| Booster 2 | |

**Phantosmia** occurs when an individual smells something that is not present. That is, there is no object that could possibly be causing the smell. For example, a person would have phantosmia if they smell smoke where there is none - and nothing could be causing the smell.

Did you experience phantosmia after your vaccination? Please answer by the dose of vaccine you received.

| | Select you answer from the drop down list |
|---|---|
| Dose 1 | |
| Dose 2 | |
| Booster | |
| Booster 2 | |

**This section is about your sense of TASTE**

How was your sense of taste before vaccination? Please answer by the dose of vaccine you received.

| | Select your answer from the drop down list |
|---|---|
| Dose 1 | |
| Dose 2 | |
| Booster | |
| Booster 2 | |

If you selected other, feel free to use this space to share more information.

Did the vaccination affect your sense of taste? Please answer by the dose of vaccine you received.

| | Yes, sense worsened | Yes, sense improved | No | Not applicable |
|---|---|---|---|---|
| Dose 1 | ○ | ○ | ○ | ○ |
| Dose 2 | ○ | ○ | ○ | ○ |

| | Yes, sense worsened | Yes, sense improved | No | Not applicable |
|---|---|---|---|---|
| Booster | ○ | ○ | ○ | ○ |
| Booster 2 | ○ | ○ | ○ | ○ |

If vaccination affected your sense of taste, when did the change start? Please answer by the dose of vaccine you received.

| | Immediately after vaccination | Within 1 day of vaccination | After 1 day of vaccination | The vaccination did not affect my sense of taste |
|---|---|---|---|---|
| Dose 1 | ○ | ○ | ○ | ○ |
| Dose 2 | ○ | ○ | ○ | ○ |
| Booster | ○ | ○ | ○ | ○ |
| Booster 2 | ○ | ○ | ○ | ○ |

If vaccination affected your sense of taste, how long did the change last? Please answer by the dose of vaccine you received.

| | <1 day | 1 day to 1 week | 1 week to 1 month | >1 month | No effect |
|---|---|---|---|---|---|
| Dose 1 | ○ | ○ | ○ | ○ | ○ |
| Dose 2 | ○ | ○ | ○ | ○ | ○ |
| Booster | ○ | ○ | ○ | ○ | ○ |
| Booster 2 | ○ | ○ | ○ | ○ | ○ |

If vaccination affected your sense of taste, which senses were affected? Please answer by the dose of vaccine you receive.

| | Please answer by the dose of vaccine | | | |
|---|---|---|---|---|
| | Dose 1 | Dose 2 | Booster | Booster 2 |
| Sweetness | ☐ | ☐ | ☐ | ☐ |
| Saltiness | ☐ | ☐ | ☐ | ☐ |
| Sourness | ☐ | ☐ | ☐ | ☐ |
| Bitterness | ☐ | ☐ | ☐ | ☐ |
| Savoriness | ☐ | ☐ | ☐ | ☐ |
| Other (Please use the space below to specify) | ☐ | ☐ | ☐ | ☐ |

If you selected other, feel free to use this space to share more information.

About distorted taste, i.e., things taste different than they did before: (Please answer by the dose of vaccine you received)

| | Select your answer from the drop down list |
|---|---|
| Dose 1 | |
| Dose 2 | |
| Booster | |
| Booster 2 | |

**This section is about your sense of CHEMESTHESIS (spicy, hot feeling by for example wasabi, chili pepper, sense of coolness by mint)**

Did the vaccination affect your sense of chemesthesis? Please answer by the dose of vaccine you received.

| | Yes, sense worsened | Yes, sense improved | No | Not applicable |
|---|---|---|---|---|
| Dose 1 | ○ | ○ | ○ | ○ |
| Dose 2 | ○ | ○ | ○ | ○ |
| Booster | ○ | ○ | ○ | ○ |
| Booster 2 | ○ | ○ | ○ | ○ |

If vaccination affected your sense of chemesthesis, when did the change start? Please answer by the dose of vaccine you received.

| | Immediately after vaccination | Within 1 day of vaccination | After 1 day of vaccination | The vaccination did not affect my sense of chemesthesis |
|---|---|---|---|---|
| Dose 1 | ○ | ○ | ○ | ○ |
| Dose 2 | ○ | ○ | ○ | ○ |
| Booster | ○ | ○ | ○ | ○ |
| Booster 2 | ○ | ○ | ○ | ○ |

If vaccination affected your sense of chemesthesis, how long did the change last? Please answer by the dose of vaccine you received.

| | <1 day | 1 day to 1 week | 1 week to 1 month | >1 month | No effect |
|---|---|---|---|---|---|
| Dose 1 | ○ | ○ | ○ | ○ | ○ |

| | <1 day | 1 day to 1 week | 1 week to 1 month | >1 month | No effect |
|---|---|---|---|---|---|
| Dose 2 | ○ | ○ | ○ | ○ | ○ |
| Booster | ○ | ○ | ○ | ○ | ○ |
| Booster 2 | ○ | ○ | ○ | ○ | ○ |

### COVID-19

#### Did you get COVID-19?

- ○ Yes, diagnosed with PCR, antibody test, or rapid antigen test
- ○ Not tested but I suspect that I got it
- ○ No or not aware

When did you get COVID-19? You can enter up to the first three times of infection.

| | Month | Year | Type of diagnosis |
|---|---|---|---|
| First time | ⌄ | ⌄ | ⌄ |
| Second time | ⌄ | ⌄ | ⌄ |
| Third time | ⌄ | ⌄ | ⌄ |

When did you recover from COVID-19? Please answer by the event of infection.

| | Month | Year | Type of test | I have not recovered |
|---|---|---|---|---|
| First time | ⌄ | ⌄ | ⌄ | ○ |
| Second time | ⌄ | ⌄ | ⌄ | ○ |
| Third time | ⌄ | ⌄ | ⌄ | ○ |

How do you consider the severity of your symptoms? Please answer by the event of COVID-19 infection.

| | Mild | Moderate | Severe (home care) | Severe (hospitalized) | Not applicable |
|---|---|---|---|---|---|
| First time | ○ | ○ | ○ | ○ | ○ |
| Second time | ○ | ○ | ○ | ○ | ○ |
| Third time | ○ | ○ | ○ | ○ | ○ |

Please select all the symptoms you experienced.

| | Please answer by the event of COVID-19 infection | | |
|---|---|---|---|
| | First time | Second time | Third time |
| Fever | ☐ | ☐ | ☐ |
| Headache | ☐ | ☐ | ☐ |
| Migraine | ☐ | ☐ | ☐ |
| Chills | ☐ | ☐ | ☐ |
| Diarrhea | ☐ | ☐ | ☐ |
| Vomit | ☐ | ☐ | ☐ |
| Stomach ache | ☐ | ☐ | ☐ |
| Joint ache | ☐ | ☐ | ☐ |
| Cough | ☐ | ☐ | ☐ |
| Sore throat | ☐ | ☐ | ☐ |
| Shortness of breath | ☐ | ☐ | ☐ |
| Muscle soreness/pain | ☐ | ☐ | ☐ |
| Lightheadedness | ☐ | ☐ | ☐ |
| Dizziness | ☐ | ☐ | ☐ |
| Nausea | ☐ | ☐ | ☐ |
| Fatigue | ☐ | ☐ | ☐ |
| Nasal congestion | ☐ | ☐ | ☐ |
| Dry/burning nose | ☐ | ☐ | ☐ |
| Heart problems | ☐ | ☐ | ☐ |
| Sleep disturbances | ☐ | ☐ | ☐ |
| Blood clots | ☐ | ☐ | ☐ |
| Skin rash | ☐ | ☐ | ☐ |
| Menstral change | ☐ | ☐ | ☐ |
| Brain fog | ☐ | ☐ | ☐ |
| Syncope (fainting/passing out) | ☐ | ☐ | ☐ |
| Smell dysfunction | ☐ | ☐ | ☐ |
| Taste dysfunction | ☐ | ☐ | ☐ |
| Reduced ability to sense spiciness | ☐ | ☐ | ☐ |
| Other (Please use the space below to specify) | ☐ | ☐ | ☐ |
| None of above | ☐ | ☐ | ☐ |
| I do not remember | ☐ | ☐ | ☐ |

If you selected other, feel free to use this space to share more information.

Did you any aura with migraine? If so, what was the type of aura? Please answer by the event of COVID-19 infection.

| | Please answer by the event of COVID-19 infection. | | |
|---|---|---|---|
| | First time | Second time | Third time |
| I did not have aura | ☐ | ☐ | ☐ |
| Visual aura | ☐ | ☐ | ☐ |
| Auditory aura | ☐ | ☐ | ☐ |
| Olfactory aura | ☐ | ☐ | ☐ |
| Other sensory aura | ☐ | ☐ | ☐ |
| Not applicable | ☐ | ☐ | ☐ |

Did you have pneumonia while having COVID-19? Please answer by the event of COVID-19 infection.

| | Yes | No | Not applicable |
|---|---|---|---|
| First time | ○ | ○ | ○ |
| Second time | ○ | ○ | ○ |
| Third time | ○ | ○ | ○ |

Did you experience an abnormal sense of smell when you were infected with COVID-19? Please answer by the event of COVID-19 infection.

| | Yes | No | Not applicable |
|---|---|---|---|
| First time | ○ | ○ | ○ |
| Second time | ○ | ○ | ○ |
| Third time | ○ | ○ | ○ |

You told us you experienced an abnormal sense of smell due to COVID-19. Have you been undergoing medical treatments, taking supplements, trying home remedies, olfactory training, or other practices to help you recover your sense of smell?

| | Please answer by the event of COVID-19 infection | | | |
|---|---|---|---|---|
| | First time | Second time | Third time | Please specify |
| Medical treatments | ☐ | ☐ | ☐ | |
| Taking supplements | ☐ | ☐ | ☐ | |

| | Please answer by the event of COVID-19 infection | | | |
|---|---|---|---|---|
| | First time | Second time | Third time | Please specify |
| Home remedies | ☐ | ☐ | ☐ | |
| Olfactory training | ☐ | ☐ | ☐ | |
| Other (Please specify) | ☐ | ☐ | ☐ | |
| No | ☐ | ☐ | ☐ | |
| Not applicable | ☐ | ☐ | ☐ | |

Did you have a distorted sense of smell (parosmia) and/or smell odors that weren't present after COVID-19 infection?

| | Please answer by the event of COVID-19 infection. | | |
|---|---|---|---|
| | First time | Second time | Third time |
| I had a distorted smell | ☐ | ☐ | ☐ |
| I smelled odors that did not exist | ☐ | ☐ | ☐ |
| None of above | ☐ | ☐ | ☐ |
| Not applicable | ☐ | ☐ | ☐ |

Did you experience an abnormal sense of taste when you were infected with COVID-19? Please answer by the event of COVID-19 infection.

| | Yes | No | Not applicable |
|---|---|---|---|
| First time | ○ | ○ | ○ |
| Second time | ○ | ○ | ○ |
| Third time | ○ | ○ | ○ |

Which sense of taste was affected?

| | Please answer by the event of COVID-19 | | | |
|---|---|---|---|---|
| | First time | Second time | Third time | Please specify |
| Sweetness | ☐ | ☐ | ☐ | |
| Saltiness | ☐ | ☐ | ☐ | |
| Sourness | ☐ | ☐ | ☐ | |
| Bitterness | ☐ | ☐ | ☐ | |
| Savoriness | ☐ | ☐ | ☐ | |

| | Please answer by the event of COVID-19 | | | |
|---|---|---|---|---|
| | First time | Second time | Third time | Please specify |
| Other (Please specify) | ☐ | ☐ | ☐ | |
| I had distorted taste | ☐ | ☐ | ☐ | |
| I had metallic/chemical taste (Please specify whether it was in mouth or nose) | ☐ | ☐ | ☐ | |
| Not applicable | ☐ | ☐ | ☐ | |

You told us you experienced an abnormal sense of taste due to COVID-19. Have you been undergoing medical treatments, taking supplements, trying home remedies, olfactory training, or other practices to help you recover your sense of taste?

| | Please answer by the event of COVID-19 infection | | | |
|---|---|---|---|---|
| | First time | Second time | Third time | Please specify |
| Medical treatments | ☐ | ☐ | ☐ | |
| Taking supplements | ☐ | ☐ | ☐ | |
| Home remedies | ☐ | ☐ | ☐ | |
| Taste training | ☐ | ☐ | ☐ | |
| Other (please specify) | ☐ | ☐ | ☐ | |
| No | ☐ | ☐ | ☐ | |
| Not applicable | ☐ | ☐ | ☐ | |

If you received a COVID-19 vaccination **AFTER** a COVID-19 infection, did any subsided symptoms of COVID-19 reoccur?

- ○ I did not receive any COVID-19 vaccinations after my COVID-19 infection
- ○ Yes (please describe in the space below)
- ○ No

If you selected other, feel free to use this space to share more information.

Did vaccination make any difference in persisting COVID-19 symptoms?

- ○ Yes, worsened

- Yes, improved
- No
- Other (for example, worsened but turned better later, vice versa; please use the space below to specify)

If you selected other, feel free to use this space to share more information.

**About yourself**

**About yourself**

How old are you?

- 18-24 years old
- 25-34 years old
- 35-44 years old
- 45-54 years old
- 55-64 years old
- 65+ years old

In which country do you currently reside?

There are data suggesting that gender and sexual orientation may cause different health outcomes. Answering these questions is optional.

Your height and current weight.

Height (cm)

Weight (kg)

**RACE**

Explanation for race/ethnicity focus: We are interested in whether you are a member of your society's dominant or marginalized group because health, wealth, and environmental disparities are associated with minoritized race and have been a factor in health outcomes related to COVID-19. Ethnicity, ethnic group, and race are used to describe social status which varies across cultures and may involve the marginalization of indigenous peoples, descendants of enslaved Africans, and immigrants from places that do not align with the dominant group (e.g., those not of European descent in the US and Europe and Canada).

Do you consider yourself to be part of a minority ethnic or racial group in the country where you currently live?

- Yes
- No
- Unsure

**Household income**
Explanation of household income (from any member contributing to household maintenance): Household income is receiving money in exchange for labor or services, from the sale of goods or property, or as profit from financial investments. Income does not include government assistance (for example, disability, unemployment, welfare). Example: annual salary with a renewal contract, short-term contract work, gig work, part-time employment outside the home, selling goods at markets or online markets.

Do you or your household have an income?

- Yes
- No

How does your household income compare to the general population (regardless of race or ethnicity group) in the country where you currently live?

What is your smoking status?

- I never smoke
- I quitted smoking before having COVID-19
- I quitted smoking after having COVID-19
- I smoke
- Prefer not to say

Are you pregnant or were you pregnant after the outbreak of COVID-19?

- Yes
- No
- Prefer not to say

Do you have below pre-existing conditions before COVID and/or before vaccination?

- Neurological pathologies
- Psychiatric pathologies
- Head and neck trauma
- Head and neck radiation
- Nasal sinus surgery
- Impaired smell and taste perception
- Allergy
- Allergic rhinitis (Hey fever)
- Migraine without aura
- Migraine with aura
- None of above

Did you routinely take any medicine before infected COVID-19 or vaccination?

- Yes (Please use the space below to specify)
- No

If you selected Yes, feel free to use this space to share more information.

Are you a healthcare professional?

- Yes
- No

**Your journey**

We would like to invite you to include any additional information that you think would help us understand the symptomatology of COVID-19 and the influence of COVID-19 vaccine. This could include information on symptoms, treatment, emotional or physical well-being, or anything else you think has been particularly important in your journey.

Is there anything else you want to share with us?

Powered by Qualtrics
